# Alkaline phosphatase variability predicts new onset heart failure, cardiovascular mortality, and all-cause mortality in patients with type-2 diabetes mellitus: a population-based study

**DOI:** 10.1101/2021.11.27.21266928

**Authors:** Jiandong Zhou, Sharen Lee, Govinda Adhikari, Wing Tak Wong, Khalid Bin Waleed, Tong Liu, Ian Chi Kei Wong, Bernard Man Yung Cheung, Gary Tse, Qingpeng Zhang

**Affiliations:** Nuffield Department of Medicine, University of Oxford, Oxford, United Kingdom; Diabetes Research Unit, Cardiovascular Analytics Group, Hong Kong, China-UK Collaboration; Internal Medicine Department, McLaren Flint, Flint, USA; School of Life Sciences, Chinese University of Hong Kong, Hong Kong, China; Department of Cardiology, St. George’s University Hospitals Foundation NHS Trust, London, United Kingdom; Tianjin Key Laboratory of Ionic-Molecular Function of Cardiovascular disease, Department of Cardiology, Tianjin Institute of Cardiology, Second Hospital of Tianjin Medical University, Tianjin 300211, China; Department of Pharmacology and Pharmacy, University of Hong Kong, Pokfulam, Hong Kong, China; Medicines Optimisation Research and Education (CMORE), UCL School of Pharmacy, London, United Kingdom; Division of Clinical Pharmacology, Department of Medicine, University of Hong Kong, Hong Kong, China; Kent and Medway Medical School, Canterbury, United Kingdom; School of Data Science, City University of Hong Kong, Hong Kong, China

## Abstract

**Objective:** To investigate the associations of alkaline phosphatase (ALP) variability measures with new onset heart failure, cardiovascular mortality, and all-cause mortality in type 2 diabetes mellitus patients with a populational-cohort study.

**Method:** This study included patients with type 2 diabetes mellitus who presented to ambulatory, outpatient and inpatient facilities managed by the public sector in Hong Kong between January 1^st^, 2000 to December 31^st^, 2019. Comprehensive clinical and medical data including demographics, past comorbidities, medications, and laboratory examinations of complete blood, lipid/glycemic profile and their variability were collected. ALP and its variability measures were extracted. Univariable and multiple multivariable Cox regression were used to identify the associations of alkaline phosphatase variability with new onset heart failure and mortality risks. Patients were stratified into three subgroups based on the tertiles of baseline ALP level.

**Results:** The study cohort consisted of 14289 patients (52.52% males, mean age at initial drug exposure: 74.55 years old [standard deviation (SD): 12.7]). Over a mean follow up of 2513 days [interquartile range (IQR): 1151-4173]), 10182 patients suffered from all-cause mortality (incidence rate [IR]: 71.25%), 1966 patients (IR: 13.75%) died from cardiovascular causes, and 1171 patients (IR: 8.19%) developed with new onset heart failure. Higher cumulative incidences of all three outcomes were observed for the highest tertile of ALP compared to medium/low tertiles. ALP baseline and variability level predicted new onset heart failure, cardiovascular and all-cause mortality before adjusting for subclinical biomarkers (p < 0.01). Amongst the measures of ALP variability, the hazard ratio (HR) of coefficient of variation (CV) was markedly raised in particular (new onset heart failure: HR=2.73, 95% confidence interval [CI]= [1.71-4.37], p <0.0001; all-cause mortality: HR= 5.83, 95% CI= [5.01-6.79], p <0.0001; cardiovascular mortality: HR= 4.81, 95% CI= [3.36-6.88], p <0.0001).

**Conclusions:** Raised ALP level and variability are associated with increased risks of all-cause mortality, cardiovascular mortality and new onset heart failure amongst patients with type 2 diabetes mellitus.

## Introduction

Alkaline phosphatase (ALP) is a type of enzyme found in the liver, bone, bile duct, kidney, intestinal mucosa, and placenta. These enzymes are broadly classified into two types: tissue-specific (placental, germ cell, intestinal type) and non-specific (TNAP) that is present in bone, kidneys, central nervous system [1]. The levels of ALP vary with age, gender, and weight. Raised ALP level is independently associated with increased risk of adverse outcomes in patients with chronic renal disease, presumed to be mediated by vascular calcification [2, 3]. ALP hydrolyzes inorganic pyrophosphate, the endogenous inhibitor of calcium phosphate crystal formation and growth [3, 4]. Furthermore, elevated ALP promoted vascular calcification in the in-vitro model [3]. Vascular calcification is an active process that promotes atherosclerosis, arterial stiffness, and aging [5]. Thus, it highlights the importance of ALP as an essential molecular marker of vascular calcification.

Although most studies have focused on hemodialysis patients, some studies have shown that higher ALP levels are linked with adverse outcomes among survivors of stroke and myocardial infarction in the general population [2, 6]. ALP has been shown to be an independent predictor of mortality, myocardial infarction, and stent-thrombosis in patients after percutaneous coronary intervention and drug eluding stent [5]. However, it is unknown whether ALP is associated with adverse outcomes among patients with type 2 diabetes mellitus. Therefore, the present study aims to evaluate the predictive value of ALP variability towards new-onset heart failure, cardiovascular and all-cause mortality amongst patients with type 2 diabetes mellitus.

## Methods

### Study design and population

The inclusion criteria of this study were patients with type 2 diabetes mellitus who presented to the Hong Kong Hospital Authority between January 1^st^, 2000 to December 31^st^, 2019. Patients with fewer than three ALP measurements before the diagnosis of heart failure, death within 90 days of the type 2 diabetes mellitus diagnosis, human immunodeficiency viral infection and a prior diagnosis of heart failure were excluded.

The patients were identified from the Clinical Data Analysis and Reporting System (CDARS), a territory-wide database that centralizes patient information from individual local hospitals to establish comprehensive clinical and medical data, including demographics, past comorbidities, medications, and laboratory examinations. The system has been previously used by our team to examine the predictive values of visit-to-visit variability in various risk variables in the general [7, 8] and diabetes-specific cohorts [9, 10]. Clinical and biochemical parameters of the population of interest were extracted from CDARS. CHA-DS-VASc score, Charlson comorbidity index and the number of prior comorbidities were also calculated. Standard International Classification of Disease, Ninth Edition (ICD-9) codes was used to identity prior comorbidities and the study outcomes were provided in **Supplementary Table 1**. Mortality was recorded using the International Classification of Diseases, Tenth Edition (ICD-10) coding. The standard deviations (SD) of lipid and glucose profiles including fast blood glucose, HbA1c, total cholesterol (TC), triglyceride (TG), low-density lipoprotein (LDL), high-density lipoprotein (HDL) were calculated. Subclinical biomarkers were also calculated, including neutrophil-to-lymphocyte ratio as neutrophil (x10^9/L)/lymphocyte (x10^9/L), albumin-to-ALP as albumin (g/L)/ALP (U/L), LDL-HDL ratio as LDL (mmol/L)/HDL (mmol/L), TC-HDL ratio as TC (mmol/L)/HDL (mmol/L), and TG-HDL ratio as TG (mmol/L)/HDL (mmol/L). The SD, variance, coefficient of variation (CV) and root mean square (RMS) of ALP were calculated. Detailed definitions about the calculations of these variability measures were provided in **Supplementary Table 2**.

### Statistical analysis and primary outcomes

The study outcomes were new onset heart failure, cardiovascular mortality (ICD-10 codes I00-I09, I11, I13, I20-I51) and all-cause mortality. Patients were followed up to the endpoints of new onset heart failure, mortality, or the end of the study (December 31^st^, 2019). Mortality data were obtained from the Hong Kong Death Registry, a population-based official government registry with the registered death records of all Hong Kong citizens. There was no adjudication of the outcomes as this relied on the ICD-9 coding or a record in the death registry. However, the coding was performed by the clinicians or administrative staff, who were not involved in the mode development.

Descriptive statistics are used to summarize baseline clinical characteristics of the study cohort. Continuous variables were presented as median (95% confidence interval [CI] or interquartile range [IQR]) and categorical variables were presented as count (%). The Mann-Whitney U test was used to compare continuous variables. The χ2 test with Yates’ correction was used for 2×2 contingency data. Univariable logistic regression identifies significant mortality risk predictors. Odds ratios (ORs) with corresponding 95% CIs and P-values were reported. There was no imputation performed for missing data. Univariable and multiple multivariable adjustment models were used to identify the associations of ALP variability with new onset heart failure and mortality risks. All significance tests were two-tailed and considered significant if P value < 0.05. There was no imputation performed for missing data. No blinding was performed for the predictor as the values were obtained from the electronic health records automatically. Data analyses were performed using RStudio software (Version: 1.1.456) and Python (Version: 3.6).

## Results

### Basic characteristics

The study cohort consists of 14289 patients (52.52% males, mean age at initial drug exposure: 74.55 years old [SD: 12.7]) after excluding 10464 patients with less than three ALP measurements before the diagnosis of type 2 diabetes mellitus (N=9488), death within 90 days of type 2 diabetes mellitus diagnosis (N=264), human immunodeficiency viral infection (N=20), with a history of heart failure (N=376), and at least one of the three ALP measurements were performed after new onset heart failure (N=316) (**Figure 1**). Over a mean follow up duration: 2513 days [IQR: 1151-4173]), 10182 patients died (incidence rate [IR]: 71.25%), 1966 patients (IR: 13.75%) died from cardiovascular causes, and 1171 patients (IR: 8.19%) developed new onset heart failure.

**Figure 1.**
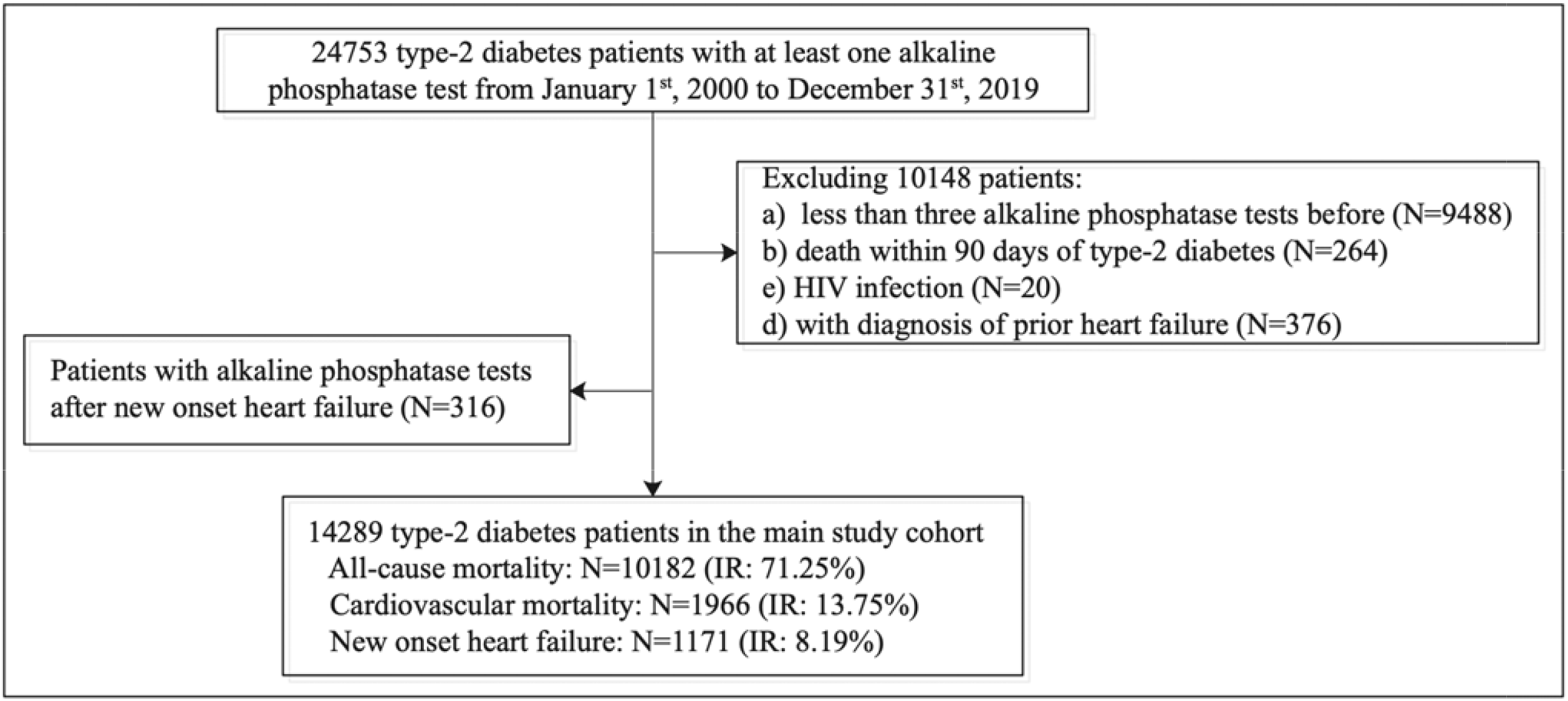
Procedures of constructing study cohorts. IR: incidence rate.

The baseline characteristics of the study cohort are summarized in **Table 1**, where patients were divided into three groups according to the tertiles of baseline ALP:

**Table 1.** Comparison of baseline and clinical characteristics of type-2 diabetes patients according to tertiles of alkaline phosphatase. * for p≤ 0.05, ** for p ≤ 0.01, *** for p ≤ 0.001, # indicates the difference among three tertiles, ACEI: angiotensin-converting-enzyme inhibitors, ARB: angiotensin II receptor blockers, CV: cardiovascular, LDL: low density lipoprotein cholesterol, HDL: high density lipoprotein cholesterol, TC: total cholesterol, TG: triglyceride, NLR: neutrophil-to-lymphocyte, AALP: albumin-to-alkaline phosphatase, SD: standard deviation, SMD: standard mean difference.

| Characteristics | Overall (N=14289)<br>Median (IQR);N or Count(%) | Low tertile (N=4604)<br>Median (IQR);N or Count(%) | Middle tertile (N=4912)<br>Median (IQR);N or Count(%) | High tertile (N=4773)<br>Median (IQR);N or Count(%) | P value<sup>#</sup> |
| --- | --- | --- | --- | --- | --- |
| ***Number of adverse events*** |  |  |  |  |  |
| All-cause mortality | 10182(71.25%) | 3006(65.29%) | 3485(70.94%) | 3691(77.33%) | <0.0001*** |
| Cardiovascular mortality | 1966(13.75%) | 595(12.92%) | 694(14.12%) | 677(14.18%) | 0.0378* |
| New onset heart failure | 1171(8.19%) | 318(6.90%) | 394(8.02%) | 459(9.61%) | <0.0001*** |
| ***Demographics*** |  |  |  |  |  |
| Male gender | 7505(52.52%) | 2663(57.84%) | 2561(52.13%) | 2281(47.78%) | 0.0110* |
| Female gender | 6784(47.47%) | 1941(42.15%) | 2351(47.86%) | 2492(52.21%) | <0.0001*** |
| Baseline age, years | 74.55(64.92-81.06);n=14289 | 74.02(63.85-80.35);n=4604 | 75.05(65.49-81.2);n=4912 | 75.05(65.05-82.06);n=4773 | 0.0018** |
| <50 | 655(4.58%) | 264(5.73%) | 191(3.88%) | 200(4.19%) | 0.433 |
| [50-60] | 1724(12.06%) | 556(12.07%) | 570(11.60%) | 598(12.52%) | 0.0302* |
| [60-70] | 2642(18.48%) | 916(19.89%) | 874(17.79%) | 852(17.85%) | 0.8778 |
| [70-80] | 4850(33.94%) | 1591(34.55%) | 1698(34.56%) | 1561(32.70%) | 0.8996 |
| >80 | 4419(30.92%) | 1277(27.73%) | 1580(32.16%) | 1562(32.72%) | <0.0001*** |
| ***Past comorbidities*** |  |  |  |  |  |
| CHA-DS-VASc Score | 3(2-4);n=14289 | 2(1-4);n=4604 | 3(2-4);n=4912 | 3(2-4);n=4773 | <0.0001*** |
| Charlson standard comorbidity index | 4(2-5);n=14289 | 2(2-4);n=4604 | 4(3-5);n=4912 | 4(2-5);n=4773 | 0.0019** |
| Number of comorbidities | 2(1-4);n=14289 | 2(1-4);n=4604 | 2(1-4);n=4912 | 2(1-4);n=4773 | 0.0285* |
| Systemic embolism | 26(0.18%) | 4(0.08%) | 11(0.22%) | 11(0.23%) | 0.3647 |
| Hypertension | 7314(51.18%) | 2295(49.84%) | 2498(50.85%) | 2521(52.81%) | 0.0004*** |
| Coronary heart disease | 3494(24.45%) | 1190(25.84%) | 1192(24.26%) | 1112(23.29%) | 0.7924 |
| Atrial fibrillation | 780(5.45%) | 234(5.08%) | 259(5.27%) | 287(6.01%) | 0.0068** |
| Hemiplegia or paraplegia | 327(2.28%) | 91(1.97%) | 128(2.60%) | 108(2.26%) | 0.7601 |
| Renal diseases | 932(6.52%) | 277(6.01%) | 306(6.22%) | 349(7.31%) | 0.0008*** |
| Osteoporosis | 6(0.04%) | 0(0.00%) | 3(0.06%) | 3(0.06%) | 0.6149 |
| Liver diseases | 105(0.73%) | 10(0.21%) | 33(0.67%) | 62(1.29%) | <0.0001*** |
| Ventricular tachycardia/fibrillation | 160(1.11%) | 43(0.93%) | 66(1.34%) | 51(1.06%) | 0.975 |
| Dementia | 658(4.60%) | 175(3.80%) | 235(4.78%) | 248(5.19%) | 0.0030** |
| Anemia | 7436(52.04%) | 2286(49.65%) | 2512(51.14%) | 2638(55.26%) | <0.0001*** |
| Acute myocardial infarction | 572(4.00%) | 169(3.67%) | 220(4.47%) | 183(3.83%) | 0.9963 |
| Chronic obstructive pulmonary disease | 570(3.98%) | 159(3.45%) | 230(4.68%) | 181(3.79%) | 0.9085 |
| Peripheral vascular disease | 168(1.17%) | 41(0.89%) | 63(1.28%) | 64(1.34%) | 0.1144 |
| Stroke/transient ischemic attack | 1335(9.34%) | 427(9.27%) | 460(9.36%) | 448(9.38%) | 0.2798 |
| Gastrointestinal bleeding | 656(4.59%) | 192(4.17%) | 232(4.72%) | 232(4.86%) | 0.0849 |
| Cancer | 121(0.84%) | 39(0.84%) | 38(0.77%) | 44(0.92%) | 0.3635 |
| Obesity | 46(0.32%) | 16(0.34%) | 14(0.28%) | 16(0.33%) | 0.8144 |
| ***Drug prescriptions*** |  |  |  |  |  |
| Number of diabetes mellitus drugs | 1(1-1);n=14289 | 1(1-2);n=4604 | 1(1-1);n=4912 | 1(1-1);n=4773 | 0.2339 |
| Number of CV drugs | 2(1-3);n=14289 | 2(1-3);n=4604 | 2(1-3);n=4912 | 2(1-3);n=4773 | <0.0001*** |
| ACEI/ARB | 7904(55.31%) | 2766(60.07%) | 2805(57.10%) | 2333(48.87%) | <0.0001*** |
| Beta blockers | 6550(45.83%) | 2247(48.80%) | 2241(45.62%) | 2062(43.20%) | 0.3993 |
| Calcium channel blockers | 7355(51.47%) | 2186(47.48%) | 2627(53.48%) | 2542(53.25%) | 0.0002*** |
| Diuretics | 5567(38.96%) | 1722(37.40%) | 1893(38.53%) | 1952(40.89%) | <0.0001*** |
| Statins | 5221(36.53%) | 1927(41.85%) | 1816(36.97%) | 1478(30.96%) | <0.0001*** |
| Metformin | 2503(17.51%) | 962(20.89%) | 819(16.67%) | 722(15.12%) | 0.0014** |
| Sulfadiazine | 12176(85.21%) | 3918(85.09%) | 4209(85.68%) | 4049(84.83%) | 0.0408* |
| Insulin | 1682(11.77%) | 431(9.36%) | 571(11.62%) | 680(14.24%) | <0.0001*** |
| Sulphonylurea | 3009(21.05%) | 1052(22.84%) | 982(19.99%) | 975(20.42%) | 0.7411 |
| Gliclazide | 10377(72.62%) | 3211(69.74%) | 3648(74.26%) | 3518(73.70%) | 0.0025** |
| Meglitinide | 893(6.24%) | 301(6.53%) | 303(6.16%) | 289(6.05%) | 0.8868 |
| Alpha-glucosidase inhibitors | 315(2.20%) | 98(2.12%) | 104(2.11%) | 113(2.36%) | 0.1715 |
| Other anti-dabetic drugs | 12588(88.09%) | 4044(87.83%) | 4340(88.35%) | 4204(88.07%) | 0.0206* |
| ***Subclinical biomarkers*** |  |  |  |  |  |
| NLR ratio | 3.17(2.11-5.18);n=11905 | 2.95(2.02-4.92);n=3731 | 3.19(2.11-5.2);n=4058 | 3.31(2.2-5.41);n=4116 | <0.0001*** |
| AALP ratio | 0.45(0.34-0.57);n=14289 | 0.64(0.55-0.74);n=4604 | 0.46(0.4-0.51);n=4912 | 0.3(0.23-0.36);n=4773 | <0.0001*** |
| LDL/HDL ratio | 2.44(1.79-3.22);n=6029 | 2.43(1.81-3.16);n=2123 | 2.48(1.8-3.25);n=2104 | 2.42(1.74-3.23);n=1802 | 0.3139 |
| TC/HDL ratio | 3.96(3.17-4.95);n=11177 | 3.97(3.17-4.92);n=3736 | 4.01(3.22-4.97);n=3884 | 3.87(3.12-4.95);n=3557 | 0.0214* |
| TG/HDL ratio | 1.21(0.76-1.98);n=11177 | 1.19(0.75-1.94);n=3736 | 1.23(0.77-2);n=3884 | 1.21(0.75-2);n=3557 | 0.798 |
| ***Complete blood counts*** |  |  |  |  |  |
| Lymphocyte, x10<sup>9</sup>/L | 1.57(1.1-2.04);n=11905 | 1.6(1.2-2.06);n=3731 | 1.59(1.1-2.08);n=4058 | 1.5(1.1-2);n=4116 | <0.0001*** |
| Neutrophil, x10<sup>9</sup>/L | 5(3.74-6.8);n=11905 | 4.8(3.6-6.54);n=3731 | 5.1(3.8-6.9);n=4058 | 5.12(3.8-7);n=4116 | 0.0039** |
| Potassium, mmol/L | 4.2(3.8-4.52);n=14211 | 4.2(3.8-4.57);n=4572 | 4.2(3.8-4.53);n=4896 | 4.16(3.8-4.5);n=4743 | 0.0094** |
| Albumin, g/L | 38(33.79-42);n=14289 | 39(34.7-42.48);n=4604 | 38(34-42);n=4912 | 37(32.4-41);n=4773 | <0.0001*** |
| Sodium, mmol/L | 139.86(137-141.9);n=14211 | 140(137.61-141.88);n=4572 | 139.89(137-142);n=4896 | 139.2(137-141.65);n=4743 | <0.0001*** |
| Urea, mmol/L | 7.02(5.2-10.6);n=14170 | 7.1(5.3-10.2);n=4562 | 7(5.2-10.4);n=4880 | 7.1(5-11.2);n=4728 | 0.7442 |
| Creatinine, umol/L | 102(77-149);n=14215 | 102(79-144);n=4573 | 101(77-148);n=4897 | 102(74-156);n=4745 | 0.8522 |
| Total protein, g/L | 73.4(68-78);n=14281 | 73.41(68-78);n=4603 | 73.65(68-78.13);n=4910 | 73(68-78.26);n=4768 | 0.4277 |
| Alkaline phosphatase, U/L | 83(67-104);n=13884 | 63(55-74);n=4458 | 82(72-94);n=4759 | 108(91-133);n=4667 | <0.0001*** |
| Aspartate transaminase, U/L | 24(18-33);n=5106 | 23(18-30);n=1631 | 23(18-30);n=1771 | 27(19.5-42.5);n=1704 | <0.0001*** |
| Alanine transaminase, U/L | 19(14-29);n=12125 | 20(14-29);n=3824 | 19(13-29);n=4136 | 20(14-31);n=4165 | 0.0036** |
| Bilirubin, umol/L | 11(7.64-15);n=14280 | 11(8-15);n=4603 | 10.5(7.07-14.7);n=4909 | 11(7.31-16);n=4768 | 0.004** |
| ***Lipid/glycemic profile and variability*** |  |  |  |  |  |
| Fasting glucose, mmol/L | 7.62(6.2-10.3);n=13848 | 7.5(6.1-9.72);n=4470 | 7.6(6.19-10.2);n=4746 | 7.8(6.3-11);n=4632 | <0.0001*** |
| SD of fasting glucose | 1.35(0.83-2.16);n=9516 | 1.23(0.77-1.96);n=3224 | 1.34(0.81-2.15);n=3267 | 1.52(0.92-2.42);n=3025 | <0.0001*** |
| HbA1c, g/dL | 12(10.5-13.5);n=13160 | 12.2(10.7-13.6);n=4164 | 12.1(10.6-13.5);n=4513 | 11.8(10.3-13.4);n=4483 | <0.0001*** |
| SD of HbA1c | 0.69(0.43-1.15);n=9741 | 0.64(0.4-1.05);n=3230 | 0.68(0.42-1.13);n=3325 | 0.77(0.47-1.31);n=3186 | <0.0001*** |
| LDL, mmol/L | 2.73(2.17-3.4);n=6624 | 2.75(2.19-3.39);n=2267 | 2.75(2.19-3.43);n=2304 | 2.7(2.14-3.38);n=2053 | 0.0289* |
| SD of LDL | 0.55(0.32-0.86);n=2635 | 0.52(0.31-0.83);n=986 | 0.55(0.32-0.85);n=929 | 0.57(0.33-0.89);n=720 | 0.1505 |
| HDL, mmol/L | 1.13(0.93-1.38);n=11177 | 1.12(0.93-1.37);n=3736 | 1.13(0.93-1.38);n=3884 | 1.13(0.92-1.4);n=3557 | 0.7603 |
| SD of HDL | 0.15(0.1-0.21);n=6885 | 0.13(0.09-0.19);n=2444 | 0.15(0.1-0.21);n=2387 | 0.16(0.1-0.23);n=2054 | <0.0001*** |
| TC, mmol/L | 4.51(3.84-5.3);n=12190 | 4.5(3.84-5.23);n=4004 | 4.59(3.9-5.33);n=4219 | 4.5(3.79-5.3);n=3967 | 0.0079** |
| SD of TC | 0.63(0.4-0.95);n=8144 | 0.62(0.4-0.94);n=2856 | 0.61(0.4-0.92);n=2828 | 0.66(0.42-0.98);n=2460 | <0.0001*** |
| TG, mmol/L | 1.35(0.97-1.94);n=12155 | 1.32(0.96-1.91);n=3994 | 1.37(0.98-1.96);n=4204 | 1.36(0.96-1.95);n=3957 | 0.6992 |
| SD of TG | 0.41(0.24-0.71);n=8096 | 0.4(0.23-0.7);n=2844 | 0.41(0.24-0.7);n=2813 | 0.42(0.25-0.74);n=2439 | 0.0158* |
| ***Alkaline phosphatase variability measures*** |  |  |  |  |  |
| Baseline, U/L | 83(67-104);n=14289 | 61(54-67);n=4604 | 82(77-89);n=4912 | 117(104-143);n=4773 | <0.0001*** |
| Mean, U/L | 83.56(68.43-104.67);n=14289 | 62.75(55.33-70.27);n=4604 | 82.5(75.5-90.67);n=4912 | 113.33(99.5-138);n=4773 | <0.0001*** |
| Median, U/L | 82.5(67.5-103);n=14289 | 62.5(55-69.5);n=4604 | 82(75-90);n=4912 | 112(98.5-135.5);n=4773 | <0.0001*** |
| Variance | 84.5(24.5-265.48);n=14289 | 38.12(12.33-112.42);n=4604 | 72(24.5-180.5);n=4912 | 227.81(80.92-665.65);n=4773 | <0.0001*** |
| SD | 9.19(4.95-16.29);n=14289 | 6.17(3.51-10.6);n=4604 | 8.49(4.95-13.44);n=4912 | 15.09(9-25.8);n=4773 | <0.0001*** |
| RMS | 84.3(68.84-105.86);n=14289 | 63.03(55.58-70.6);n=4604 | 82.95(75.77-91.14);n=4912 | 114.38(100.14-140.81);n=4773 | <0.0001*** |
| CV | 0.09(0.05-0.16);n=14289 | 0.09(0.05-0.14);n=4604 | 0.09(0.05-0.14);n=4912 | 0.12(0.07-0.19);n=4773 | <0.0001*** |

a. Low tertile: ALP <72 U/L;
b. Middle tertile: ALP ≥72 U/L and ≤90 U/L;
c. High tertile: ALP>90 U/L.

The baseline and clinical characteristics of patients with new onset heart failure and mortality are summarized in **Supplementary Table 3**. Kaplan Meier survival curves and cumulative incidence curves of new onset heart failure, cardiovascular mortality, and all-cause mortality stratified by high tertile v.s. low or middle tertile of ALP respectively is shown in **Figure 2 and 3**. The cumulative incidence for cardiovascular and all-cause mortality and new onset heart failure was significantly higher in the higher tertile of ALP compared to medium/low tertiles.

**Figure 2.**
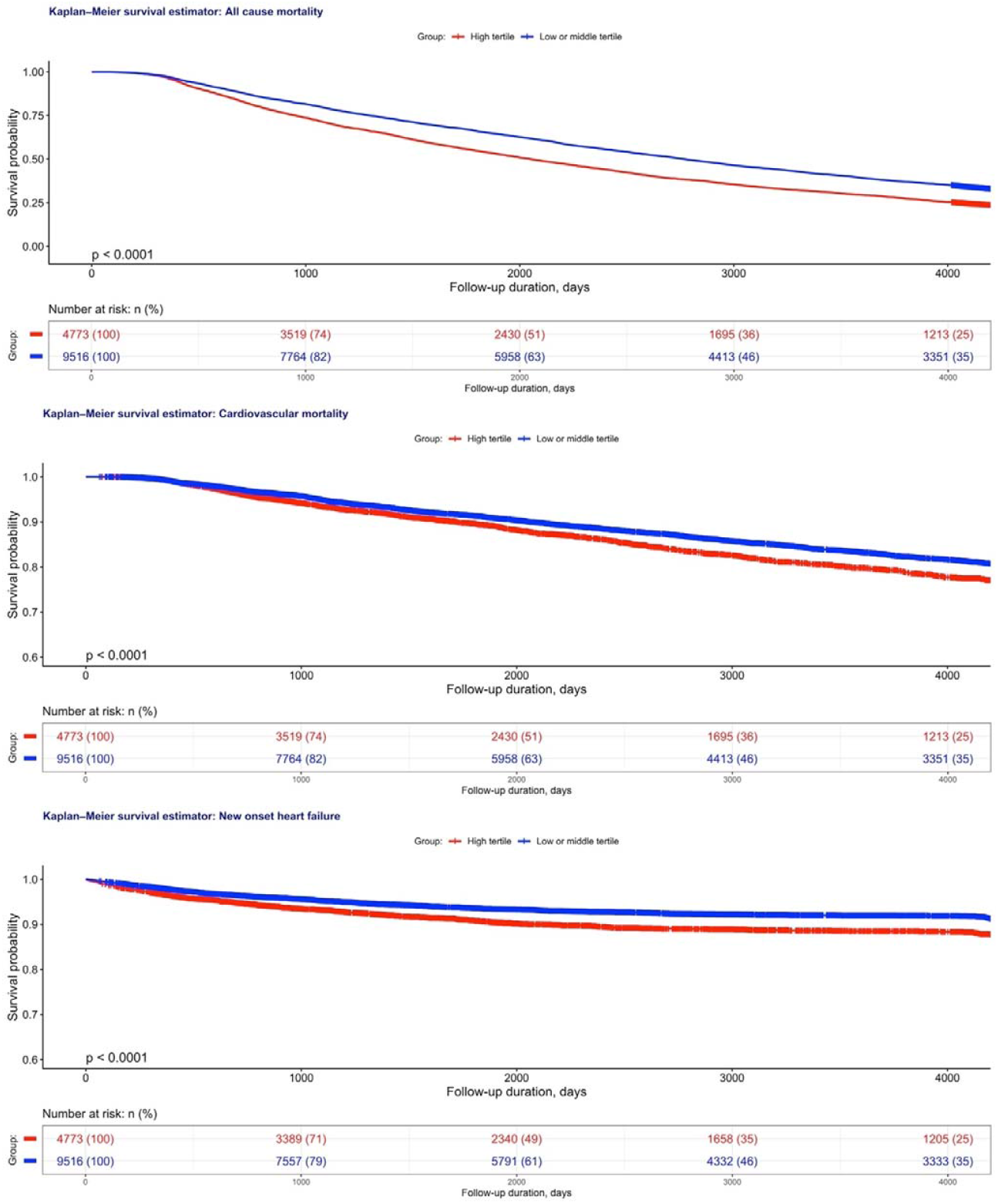
Kaplan Meier survival curves of all-cause mortality, cardiovascular mortality and new onset heart failure stratified by high tertile v.s. low or middle tertile of alkaline phosphatase.

**Figure 3.**
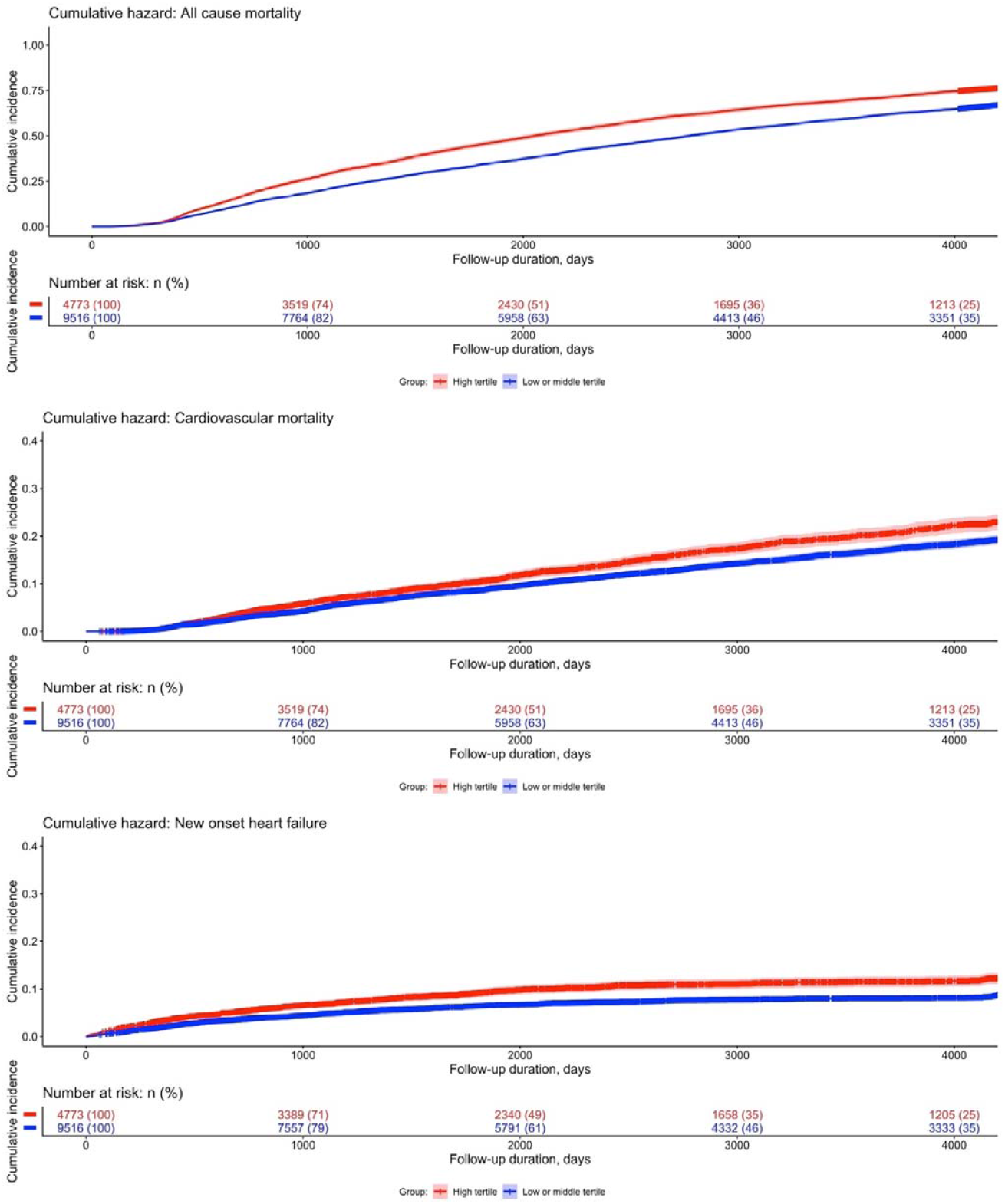
Cumulative curves of all-cause mortality, cardiovascular mortality and new onset heart failure stratified by high tertile v.s. low or middle tertile of alkaline phosphatase.

### Subgroup analysis: interactions between age and high v.s. low or middle tertile of alkaline phosphatase

Kaplan-Meier curves and cumulative incidence curves of new onset heart failure, cardiovascular mortality, and all-cause mortality among type 2 diabetes mellitus patients stratified by age and tertiles of ALP are shown in **Figures 4 and 5**. Survival against adverse outcomes differed significantly across different age groups (p < 0.0001). There was a trend towards increasing cumulative hazard in the older age group (age ≥ 75 years) across all endpoints. The cumulative risk was higher in the older medium/low tertile group (≥ 75 years, ALP < 90 IU/L) compared to the relatively younger high tertile group (< 75 years, ALP ≥ 90 U/L).

**Figure 4.**
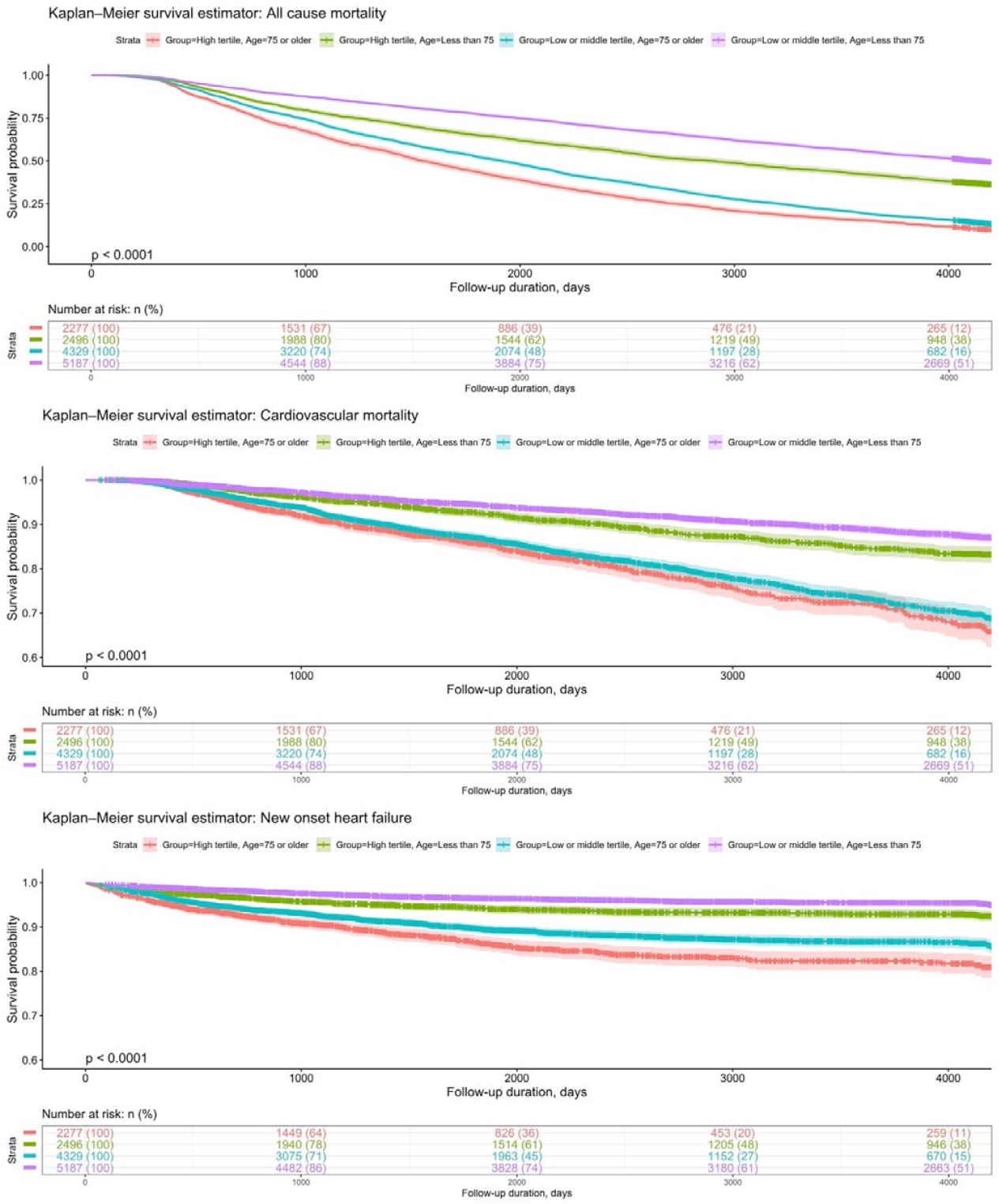
Kaplan-Meier curves of all-cause mortality, cardiovascular mortality and new onset heart failure among type-2 diabetes mellitus patients stratified by interactions formed by age and high v.s. low or middle tertile of alkaline phosphatase.

**Figure 5.**
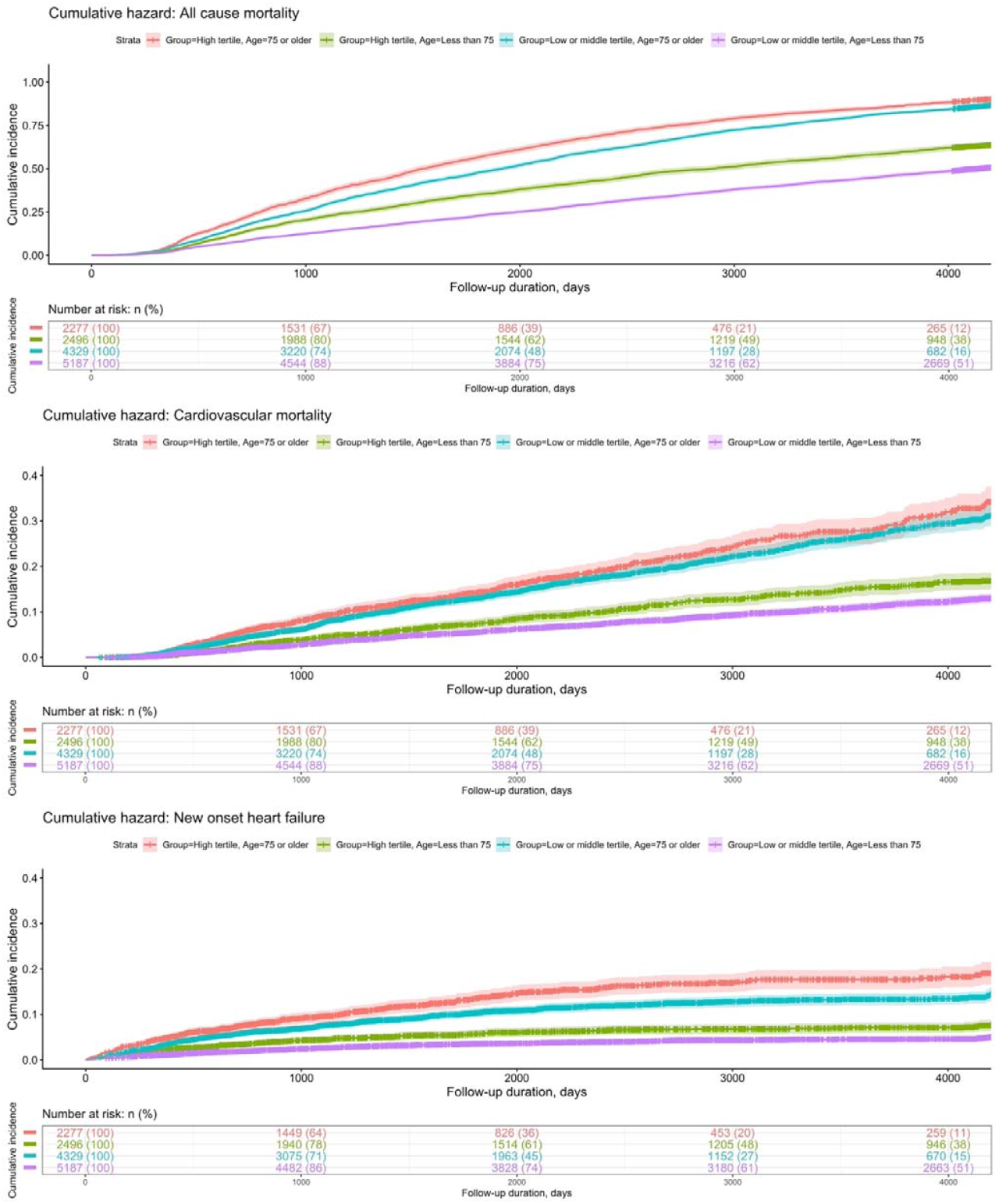
Cumulative incidence curves of all-cause mortality, cardiovascular mortality and new onset heart failure among type-2 diabetes mellitus patients stratified by interactions formed by age and high v.s. low or middle tertile of alkaline phosphatase.

### Cox regression models

Univariable Cox regression was applied to identify significant predictors of new onset heart failure, cardiovascular mortality, and all-cause mortality (**Supplementary Table 4**). Multivariable analysis adjusted for confounding variables were then conducted (**Table 2**). ALP baseline and variability level were predictive of new onset heart failure, cardiovascular and all-cause mortality before adjusting for subclinical biomarkers (p < 0.01). Amongst the variability measures, the HR of CV was markedly raised in particular (new onset heart failure: HR=2.73, 95% CI= [1.71-4.37], p <0.0001; all-cause mortality: HR= 5.83, 95% CI= [5.01-6.79], p <0.0001; cardiovascular mortality: HR= 4.81, 95% CI= [3.36-6.88], p <0.0001). However, after adjusting for subclinical biomarkers the statistical significance of ALP baseline and variability level for new-onset heart failure is lost.Finally, the marginal effects of ALP and its variability measures stratified by age and gender were explored (**Supplementary Figure 2A-E and 3A-E**).

**Table 2.** Multivariable Cox regression models for new onset heart failure, cardiovascular mortality, and all-cause mortality. * for p≤ 0.05, ** for p ≤ 0.01, *** for p ≤ 0.001; ACEI: angiotensin-converting-enzyme inhibitors, ARB: angiotensin II receptor blockers, CV: cardiovascular, LDL: low density lipoprotein cholesterol, HDL: high density lipoprotein cholesterol, TC: total cholesterol, TG: triglyceride, SD: standard deviation, SMD: standard mean difference.

| **Model 1: Adjusted by demographics** |  |  |  |
| --- | --- | --- | --- |
| **Characteristics** | **All-cause mortality HR [95% CI];P value** | **Cardiovascular mortality HR [95% CI];P value** | **New onset heart failure HR [95% CI];P value** |
| Baseline alkaline phosphatase, U/L | 1.002[1.002-1.002];<0.0001*** | 1.002[1.001-1.002];<0.0001*** | 1.002[1.001-1.003];<0.0001*** |
| <72 U/L | 1.0[Reference] | 1.0[Reference] | 1.0[Reference] |
| 72-90 U/L | 0.95[0.91-0.99];0.0108* | 1.00[0.91-1.09];0.9295 | 0.93[0.82-1.05];0.2372 |
| >90 U/L | 1.35[1.29-1.40];<0.0001*** | 1.22[1.11-1.34];<0.0001*** | 1.39[1.24-1.57];<0.0001*** |
| Mean of alkaline phosphatase, U/L | 1.003[1.003-1.003];<0.0001*** | 1.002[1.002-1.003];<0.0001*** | 1.002[1.001-1.003];<0.0001*** |
| Median of alkaline phosphatase, U/L | 1.003[1.002-1.003];<0.0001*** | 1.002[1.001-1.003];<0.0001*** | 1.002[1.001-1.003];<0.0001*** |
| Variance of alkaline phosphatase | 1.000[1.000-1.000];<0.0001*** | 1.000[1.000-1.000];0.3232 | 1.000[1.000-1.000];0.9483 |
| SD of alkaline phosphatase | 1.00[1.00-1.01];<0.0001*** | 1.00[1.00-1.01];<0.0001*** | 1.003[1.002-1.005];<0.0001*** |
| RMS of alkaline phosphatase | 1.003[1.002-1.003];<0.0001*** | 1.002[1.002-1.003];<0.0001*** | 1.002[1.001-1.002];<0.0001*** |
| CV of alkaline phosphatase | 6.12[5.27-7.10];<0.0001*** | 5.49[3.89-7.75];<0.0001*** | 5.76[3.77-8.79];<0.0001*** |
| **Model 2: Adjusted by demographics and past comorbidities** |  |  |  |
| **Characteristics** | **All-cause mortality HR [95% CI];P value** | **Cardiovascular mortality HR [95% CI];P value** | **New onset heart failure HR [95% CI];P value** |
| Baseline alkaline phosphatase, U/L | 1.002[1.002-1.002];<0.0001*** | 1.002[1.001-1.002];<0.0001*** | 1.002[1.001-1.003];<0.0001*** |
| <72 U/L | 1.0[Reference] | 1.0[Reference] | 1.0[Reference] |
| 72-90 U/L | 0.94[0.90-0.98];0.0036** | 0.99[0.90-1.08];0.7507 | 0.91[0.81-1.03];0.1470 |
| >90 U/L | 1.33[1.28-1.39];<0.0001*** | 1.20[1.09-1.31];0.0002*** | 1.26[1.12-1.42];0.0001*** |
| Mean of alkaline phosphatase, U/L | 1.003[1.002-1.003];<0.0001*** | 1.002[1.001-1.003];<0.0001*** | 1.002[1.001-1.003];<0.0001*** |
| Median of alkaline phosphatase, U/L | 1.003[1.002-1.003];<0.0001*** | 1.002[1.001-1.003];<0.0001*** | 1.002[1.001-1.003];<0.0001*** |
| Variance of alkaline phosphatase | 1.000[1.000-1.000];<0.0001*** | 1.000[1.000-1.000];0.3389 | 1.000[1.000-1.000];0.9780 |
| SD of alkaline phosphatase | 1.005[1.004-1.005];<0.0001*** | 1.004[1.003-1.005];<0.0001*** | 1.003[1.001-1.005];0.0008*** |

| RMS of alkaline phosphatase | 1.002[1.002-1.003];<0.0001*** | 1.002[1.001-1.003];<0.0001*** | 1.002[1.001-1.002];<0.0001*** |
| --- | --- | --- | --- |
| CV of alkaline phosphatase | 5.73[4.92-6.67];<0.0001*** | 4.81[3.37-6.87];<0.0001*** | 3.10[1.94-4.94];<0.0001*** |
| **Model 3: Adjusted by demographics, past comorbidities, and medications** |  |  |  |
| **Characteristics** | **All-cause mortality HR [95% CI];P value** | **Cardiovascular mortality HR [95% CI];P value** | **New onset heart failure HR [95% CI];P value** |
| Baseline alkaline phosphatase, U/L | 1.002[1.002-1.002];<0.0001*** | 1.002[1.001-1.003];<0.0001*** | 1.002[1.001-1.003];<0.0001*** |
| <72 U/L | 1.0[Reference] | 1.0[Reference] | 1.0[Reference] |
| 72-90 U/L | 0.94[0.90-0.98];0.0027** | 0.98[0.89-1.07];0.6259 | 0.90[0.79-1.01];0.0752 |
| >90 U/L | 1.32[1.27-1.38];<0.0001*** | 1.22[1.11-1.34];<0.0001*** | 1.35[1.20-1.52];<0.0001*** |
| Mean of alkaline phosphatase, U/L | 1.003[1.002-1.003];<0.0001*** | 1.002[1.002-1.003];<0.0001*** | 1.002[1.001-1.003];<0.0001*** |
| Median of alkaline phosphatase, U/L | 1.002[1.002-1.003];<0.0001*** | 1.002[1.001-1.003];<0.0001*** | 1.002[1.001-1.003];<0.0001*** |
| Variance of alkaline phosphatase | 1.000[1.000-1.000];<0.0001*** | 1.000[1.000-1.000];0.2702 | 1.000[1.000-1.000];0.9344 |
| SD of alkaline phosphatase | 1.005[1.004-1.005];<0.0001*** | 1.00[1.00-1.01];<0.0001*** | 1.003[1.001-1.005];0.0013** |
| RMS of alkaline phosphatase | 1.002[1.002-1.003];<0.0001*** | 1.002[1.002-1.003];<0.0001*** | 1.002[1.001-1.002];<0.0001*** |
| CV of alkaline phosphatase | 5.83[5.01-6.79];<0.0001*** | 4.81[3.36-6.88];<0.0001*** | 2.73[1.71-4.37];<0.0001*** |
| **Model 4: Adjusted by demographics, past comorbidities, medications, and subclinical biomarkers** |  |  |  |
| **Characteristics** | **All-cause mortality HR [95% CI];P value** | **Cardiovascular mortality HR [95% CI];P value** | **New onset heart failure HR [95% CI];P value** |
| Baseline alkaline phosphatase, U/L | 0.998[0.997-0.999];0.0008*** | 0.999[0.996-1.001];0.3442 | 1.001[0.999-1.004];0.2347 |
| <72 U/L | 1.0[Reference] | 1.0[Reference] | 1.0[Reference] |
| 72-90 U/L | 0.93[0.87-1.00];0.0400* | 0.96[0.82-1.11];0.5582 | 0.88[0.73-1.06];0.1911 |
| >90 U/L | 1.05[0.78-1.34];0.2311 | 1.06[0.71-1.05];0.1442 | 1.24[0.97-1.58];0.0810 |
| Mean of alkaline phosphatase, U/L | 1.000[0.999-1.001];0.7652 | 1.000[0.998-1.002];0.8385 | 1.002[0.999-1.004];0.2015 |
| Median of alkaline phosphatase, U/L | 1.000[0.999-1.001];0.5284 | 1.000[0.998-1.002];0.9859 | 1.002[0.999-1.004];0.1910 |
| Variance of alkaline phosphatase | 1.000[1.000-1.000];0.4291 | 1.000[1.000-1.000];0.8940 | 1.000[1.000-1.000];0.3917 |
| SD of alkaline phosphatase | 1.003[1.002-1.005];<0.0001*** | 1.00[1.00-1.01];0.0294* | 1.00[1.00-1.01];0.4590 |
| RMS of alkaline phosphatase | 1.000[0.999-1.001];0.4650 | 1.000[0.998-1.002];0.7092 | 1.001[0.999-1.003];0.2721 |
| CV of alkaline phosphatase | 4.18[3.16-5.53];<0.0001*** | 3.62[1.97-6.66];<0.0001*** | 1.93[0.94-3.96];0.0741 |

## Discussion

The main findings of this study are that:

i. there is an increased risk for cardiovascular and all-cause mortality, and new onset heart failure with increasing tertile of ALP in patients with type 2 diabetes mellitus, with the association being more robust with the higher tertile of ALP (≥ 90 IU/L). The results remained after adjustment to the available potential confounders from the cohort;
ii. elderly patients above the age of 75 are at a further increased risk for new-onset heart failure, cardiovascular and all-cause mortality under high ALP level;
iii. increased ALP variability is associated with an increased risk for new-onset heart failure, cardiovascular and all-cause mortality after adjustments to demographic, clinical and medication confounders.
iv. low glycemic variability is protective against new onset heart failure, cardiovascular and all-cause mortality amongst patients with type 2 diabetes mellitus.

Elevated ALP level has been linked to poor prognosis in patients with heart failure. A posthoc analysis of SURVIVE (Survival of Patients with Acute Heart Failure in Need of Intravenous Inotropic Support) trial showed that acute decompensated heart failure patients with elevated ALP levels had increased 180-day mortality compared to those with normal ALP levels [11]. Also, elevated ALP level has been shown to predict worsening renal function in patients with acute decompensated heart failure [12]. They highlighted that ALP may independently predict subclinical renal damage or may predict venous congestion and severity of right heart failure, which are the markers of worsening renal status in acute heart failure patients [12]. In one of the retrospective studies of chronic stable heart failure patients, ALP and gamma glutamyl transferase (GGT) were found to be independent predictors of death or heart transplantation after full adjustment in both the males and females[13]. Prior studies have demonstrated that elevated bilirubin is strongly associated with a worse prognosis in heart failure patients. [13, 14]. Right ventricular dysfunction is common in both Heart failure with reduced or preserved ejection fraction and is associated with poor prognosis [15-17]. Thus, ALP as a marker of hepatic congestion and Right ventricular dysfunction is a predictor of worse prognosis in acute or chronic heart failure patients. However, the association of new-onset heart failure and adverse events with increasing tertile of ALP has not been well-known in patients with type 2 diabetes mellitus. To the best of our knowledge, our study is the first to investigate the possible association of new-onset HF with higher tertiles of ALP in type 2 diabetic patients.

Elevated ALP has been linked to increased cardiovascular and all-cause mortality in various epidemiological studies in the general population or those with impaired renal function or those with coronary artery disease. A meta-analysis has reported that in patients with preserved renal function, serum ALP positively correlated with cardiovascular mortality. The risk of total mortality increases as ALP becomes elevated [18]. It was also found that the risk of total mortality with ALP was higher among Asians, males, and those with severe diabetes mellitus. Another retrospective cohort study showed that elevated baseline ALP levels in patients with coronary artery disease were associated independently with increased risk of cardiac and all-cause mortality (Ndrepepa et al., 2017). Similarly, the association of adverse outcomes with various tertiles of ALP has been described in a study of posthoc analysis of CARE (Cholesterol and Recurrent Events) trial and Third National Health and Nutrition Examination Survey (NHANES III)[19]. The posthoc analysis of the CARE trial demonstrated that the risk of death correlated with increasing tertile of ALP even after fully adjusted analysis. [19]. Moreover, the association did not change with the presence or absence of chronic kidney disease (CKD), diabetes mellitus, CRP smoking status, body mass index, or proteinuria. However, the association was more robust when combined with elevated phosphate levels. Another prospective study showed that ALP alone was associated with an increased risk of overall cardiovascular mortality over the course of follow-up for 11 years. [20].

The association between ALP and systemic inflammation may explain the increased cardiovascular risk amongst patients with raised ALP levels. TNAP catalyzes pyrophosphate (inhibitor of calcium phosphate crystal formation) to inorganic phosphate. Elevated levels of TNAP were found in animal models with medial vascular calcification [21] and have been suggested as a potential therapeutic target as vascular calcification is presumed to be a marker for sub-clinical atherosclerosis. Current evidence suggests the potential role of ALP as a marker of cardio-metabolic risk factors as evident from increasing levels of ALP in the setting of various risk factors including age, smoking, physical inactivity, elevated systolic blood pressure, diabetes, and low-density lipoprotein [20] [19]. ALP is also thought to be a marker of inflammation and showed a positive correlation with C reactive protein (CRP) in the National Health and Nutrition Examination Survey 2005-2006 [22]. The association of ALP with vascular calcification, cardio-metabolic risk factors, and systemic inflammation highlights the mechanisms that could potentially lead to adverse clinical outcomes in patients with elevated ALP.

## Limitations

There are several limitations to be noted for the present study. First, given its observational nature, the study is susceptible to observational bias with under-coding and coding errors. Moreover, cardiovascular risk factors such as smoking and sedentary lifestyle were not documented on CDARS, hence were not extracted. Also, the serum phosphorus, vitamin D, and parathyroid hormone status was not available in our cohort and were not included during multivariable analysis. In addition, echocardiographic data were unavailable hence the left ventricular ejection fraction was not known.

## Conclusions

Higher ALP levels and ALP variability are associated with increased risks of all-cause mortality, cardiovascular mortality and new onset heart failure in patients with type 2 diabetes mellitus. Further study is needed to elucidate the underlying mechanism and validate the predictive value of ALP and its variability in the cardiovascular adverse outcomes amongst patients with diabetes mellitus.

## Data Availability

All data produced in the present study are available upon reasonable request to the authors

## References

[1] A. Sebastian-Serrano, L. de Diego-Garcia, C. Martinez-Frailes, J. Avila, H. Zimmermann, J.L. Millan, M.T. Miras-Portugal, M. Diaz-Hernandez, Tissue-nonspecific Alkaline Phosphatase Regulates Purinergic Transmission in the Central Nervous System During Development and Disease, Comput Struct Biotechnol J 13 (2015) 95–100.

[2] M. Tonelli, M. Curhan G Fau-Pfeffer, F. Pfeffer M Fau - Sacks, R. Sacks F Fau - Thadhani, M.L. Thadhani R Fau - Melamed, N. Melamed Ml Fau - Wiebe, P. Wiebe N Fau - Muntner, P. Muntner, Relation between alkaline phosphatase, serum phosphate, and all-cause or cardiovascular mortality, (1524-4539 (Electronic)).

[3] R. Villa-Bellosta, X. Wang, J.L. Millán, G.R. Dubyak, W.C. O’Neill, Extracellular pyrophosphate metabolism and calcification in vascular smooth muscle, Am J Physiol Heart Circ Physiol 301(1) (2011) H61–8.

[4] K.A. Lomashvili, S. Narisawa, J.L. Millán, W.C. O’Neill, Vascular calcification is dependent on plasma levels of pyrophosphate, (1523-1755 (Electronic)).

[5] J.-B. Park, D.-y. Kang, H.-M. Yang, H.-J. Cho, K.W. Park, H.-Y. Lee, H.-J. Kang, B.-K. Koo, H.-S. Kim, Serum alkaline phosphatase is a predictor of mortality, myocardial infarction, or stent thrombosis after implantation of coronary drug-eluting stent, European Heart Journal 34(12) (2012) 920–931.

[6] W.S. Ryu, C.K. Lee Sh Fau - Kim, B.J. Kim Ck Fau - Kim, B.W. Kim Bj Fau - Yoon, B.W. Yoon, Increased serum alkaline phosphatase as a predictor of long-term mortality after stroke, (1526-632X (Electronic)).

[7] J. Zhou, H. Li, C. Chang, W.K.K. Wu, X. Wang, T. Liu, B.M.Y. Cheung, Q. Zhang, S. Lee, G. Tse, The association between blood pressure variability and hip or vertebral fracture risk: A population-based study, Bone 150 (2021) 116015.

[8] J. Zhou, S. Lee, W.T. Wong, K.S.K. Leung, R.H.K. Nam, P.S.H. Leung, Y.A. Chau, T. Liu, C. Chang, B.M.Y. Cheung, G. Tse, Q. Zhang, Gender- and Age-Specific Associations of Visit-to-Visit Blood Pressure Variability With Anxiety, Front Cardiovasc Med 8 (2021) 650852.

[9] S. Lee, J. Zhou, W.T. Wong, T. Liu, W.K.K. Wu, I.C.K. Wong, Q. Zhang, G. Tse, Glycemic and lipid variability for predicting complications and mortality in diabetes mellitus using machine learning, BMC Endocr Disord 21(1) (2021) 94.

[10] S. Lee, T. Liu, J. Zhou, Q. Zhang, W.T. Wong, G. Tse, Predictions of diabetes complications and mortality using hba1c variability: a 10-year observational cohort study, Acta Diabetol 58(2) (2021) 171–180.

[11] M. Nikolaou, J. Parissis, M.B. Yilmaz, M.F. Seronde, M. Kivikko, S. Laribi, C. Paugam-Burtz, D. Cai, P. Pohjanjousi, P.F. Laterre, N. Deye, P. Poder, A. Cohen-Solal, A. Mebazaa, Liver function abnormalities, clinical profile, and outcome in acute decompensated heart failure, Eur Heart J 34(10) (2013) 742–9.

[12] M. Yamazoe, A. Mizuno, Y. Nishi, K. Niwa, M. Isobe, Serum alkaline phosphatase as a predictor of worsening renal function in patients with acute decompensated heart failure, J Cardiol 67(5) (2016) 412–7.

[13] G. Poelzl, M. Ess, C. Mussner-Seeber, O. Pachinger, M. Frick, H. Ulmer, Liver dysfunction in chronic heart failure: prevalence, characteristics and prognostic significance, Eur J Clin Invest 42(2) (2012) 153–63.

[14] L.A. Allen, G.M. Felker, S. Pocock, J.J. McMurray, M.A. Pfeffer, K. Swedberg, D. Wang, S. Yusuf, E.L. Michelson, C.B. Granger, C. Investigators, Liver function abnormalities and outcome in patients with chronic heart failure: data from the Candesartan in Heart Failure: Assessment of Reduction in Mortality and Morbidity (CHARM) program, Eur J Heart Fail 11(2) (2009) 170–7.

[15] J.A. Borovac, D. Glavas, Z. Susilovic Grabovac, D. Supe Domic, L. Stanisic, D. D’Amario, D. Duplancic, J. Bozic, Right Ventricular Free Wall Strain and Congestive Hepatopathy in Patients with Acute Worsening of Chronic Heart Failure: A CATSTAT-HF Echo Substudy, J Clin Med 9(5) (2020).

[16] L. Bosch, C.S.P. Lam, L. Gong, S.P. Chan, D. Sim, D. Yeo, F. Jaufeerally, K.T.G. Leong, H.Y. Ong, T.P. Ng, A.M. Richards, F. Arslan, L.H. Ling, Right ventricular dysfunction in left-sided heart failure with preserved versus reduced ejection fraction, Eur J Heart Fail 19(12) (2017) 1664–1671.

[17] T.M. Gorter, E.S. Hoendermis, D.J. van Veldhuisen, A.A. Voors, C.S. Lam, B. Geelhoed, T.P. Willems, J.P. van Melle, Right ventricular dysfunction in heart failure with preserved ejection fraction: a systematic review and meta-analysis, Eur J Heart Fail 18(12) (2016) 1472–1487.

[18] J.W. Li, C. Xu, Y. Fan, Y. Wang, Y.B. Xiao, Can serum levels of alkaline phosphatase and phosphate predict cardiovascular diseases and total mortality in individuals with preserved renal function? A systemic review and meta-analysis, PLoS One 9(7) (2014) e102276.

[19] M. Tonelli, G. Curhan, M. Pfeffer, F. Sacks, R. Thadhani, M.L. Melamed, N. Wiebe, P. Muntner, Relation between alkaline phosphatase, serum phosphate, and all-cause or cardiovascular mortality, Circulation 120(18) (2009) 1784–92.

[20] S.G. Wannamethee, N. Sattar, O. Papcosta, L. Lennon, P.H. Whincup, Alkaline phosphatase, serum phosphate, and incident cardiovascular disease and total mortality in older men, Arterioscler Thromb Vasc Biol 33(5) (2013) 1070–6.

[21] R. Villa-Bellosta, J. Egido, Phosphate, pyrophosphate, and vascular calcification: a question of balance, Eur Heart J 38(23) (2017) 1801–1804.

[22] M. Webber, A. Krishnan, N.G. Thomas, B.M. Cheung, Association between serum alkaline phosphatase and C-reactive protein in the United States National Health and Nutrition Examination Survey 2005-2006, Clin Chem Lab Med 48(2) (2010) 167–73.

