## Supplementary Appendix for "Alkaline phosphatase variability predicts new onset heart failure, cardiovascular mortality, and all-cause mortality in patients with type-2 diabetes mellitus: a population-based study"

**Supplementary Table 1. ICD-9 codes for diagnoses.**

Diabetes mellitus 250	250.01	250.02	250.03	250.1	250.11	250.12	250.13	250.2	250.21	250.22	250.23	250.3	250.31	250.32	250.33	250.4	250.41	250.42	250.43	250.5	250.51	250.52	250.53	250.6	250.61	250.62	250.63	250.7	250.71	250.72	250.73	250.8	250.81	250.82	250.83	250.9	250.91	250.92	250.93

Hypertension	401	401.1	401.9	402	402.01	402.1	402.11	402.9	402.91	403	403.01	403.1	403.11	403.9	403.91	404	404.01	404.02	404.03	404.1	404.11	404.12	404.13	404.9	404.91	404.92	404.93	405	405.01	405.09	405.1	405.11	405.19	405.9	405.91	405.99	437.2

Heart failure	428	428	428.1	428.2	428.2	428.21	428.22	428.23	428.3	428.3	428.31	428.32	428.33	428.4	428.4	428.41	428.42	428.43	428.9	398.91	402.01	402.11	402.91	404.01	404.03	404.11	404.13	404.91	404.93

Hemiplegia or paraplegia 344.1	342	342	342	342.01	342.02	342.1	342.1	342.11	342.12	342.8	342.8	342.81	342.82	342.9	342.9	342.91	342.92

Atrial fibrillation	427.31	429.4

Osteoporosis 733.00 733.01 733.02 733.03 733.09 733.41 733.42 733.43 733.40 731.0 733.7 733.00

Stroke/transient ischemic attack	435	435.1	435.2	435.3	435.8	435.9	433.81	433.91	434	436	437	437.1	433.31	433.01	434.01	434.1	434.11	434.9	434.91	437.2	437.3	437.4	437.5	437.6	437.7	437.8	437.9	430	431	432	432.1	432.9

Neurologic 250.6	250.61	251.61	252.61

Anemia 281 282 283 285

Ventricular tachycardia/fibrillation 427.41 427.1

Ophthalmic 250.5	250.51	250.52	250.53

Liver diseases	456	456.1	456.2	572.2	572.3	572.4	572.8	571.4	571.5	571.6

Dementia	331.82	290	290.1	290.11	290.12	290.13	290.2	290.21	290.3	290.4	290.41	290.42	290.43	290.8	290.9	294.2	294.1	294.11	294.21	332	46.1	333.4	340	42331	331.19	294.29

Anxiety disorder 300.0 300.01 300.02 300.2 300.21 300.23 300.3

Chronic Obstructive Pulmonary Didsease	490	491	492	493	494	495	496	491.1	491.2	491.21	491.22	491.8	491.9	492.8	493.01	493.02	493.1	493.11	493.12	493.2	493.21	493.22	493.8	493.81	493.82	493.9	493.91	493.92	494.1	495.1	495.2	495.3	495.4	495.5	495.6	495.7	495.8	495.9

Acute myocardial infarction 410.x

Peripehral vascular disease	250.7	443.9	443	443.1	443.2	443.21	443.22	443.23	443.24	443.29	443.8	443.81	443.82	443.89	441	443.9	785.4	V43.4

Gastrointestinal bleeding	531	531.2	531.4	531.6	532	532.2	532.4	532.6	533	533.2	533.4	533.6	534	534.2	534.4	534.6	535.01	535.11	535.21	535.31	535.41	535.51	535.61	535.71	562.02	562.03	562.12	562.13	569.3	569.85	569.86	578	578.1	578.9

Coronary heart disease	410.01	410.02	410.1	410.11	410.12	410.2	410.21	410.22	410.3	410.31	410.32	410.4	410.41	410.42	410.5	410.51	410.52	410.6	410.61	410.62	410.7	410.71	410.72	410.8	410.81	410.82	410.9	410.91	410.92	411	411.1	411.8	411.81	411.89	413	413.1	413.9	414	414.01	414.02	414.03	414.04	414.05	414.06	414.07	414.1	414.11	414.12	414.19	414.2	414.3	414.4	414.8	414.9	410	412

Cancer 140	140.1	140.3	140.4	140.5	140.6	140.8	140.9	141	141.1	141.2	141.3	141.4	141.5	141.6	141.8	141.9	142	142.1	142.2	142.8	142.9	143	143.1	143.8	143.9	144	144.1	144.8	144.9	145	145.1	145.2	145.3	145.4	145.5	145.6	145.8	145.9	146	146.1	146.2	146.3	146.4	146.5	146.6	146.7	146.8	146.9	147	147.1	147.2	147.3	147.8	147.9	148	148.1	148.2	148.3	148.8	148.9	149	149.1	149.8	149.9	150	150.1	150.2	150.3	150.4	150.5	150.8	150.9	151	151.1	151.2	151.3	151.4	151.5	151.6	151.8	151.9	152	152.1	152.2	152.3	152.8	152.9	153	153.1	153.2	153.3	153.4	153.5	153.6	153.7	153.8	153.9	154	154.1	154.2	154.3	154.8	155	155.1	155.2	156	156.1	156.2	156.8	156.9	157	157.1	157.2	157.3	157.4	157.8	157.9	158	158.8	158.9	159	159.1	159.8	159.9	160	160.1	160.2	160.3	160.4	160.5	160.8	160.9	161	161.1	161.2	161.3	161.8	161.9	162	162.2	162.3	162.4	162.5	162.8	162.9	163	163.1	163.8	163.9	164	164.1	164.2	164.3	164.8	164.9	165	165.8	165.9	170	170.1	170.2	170.3	170.4	170.5	170.6	170.7	170.8	170.9	171	171.2	171.3	171.4	171.5	171.6	171.7	171.8	171.9	172	172.1	172.2	172.3	172.4	172.5	172.6	172.7	172.8	172.9	and	173	173.01	173.02	173.09	173.1	173.11	173.12	173.19	173.2	173.21	173.22	173.29	173.3	173.31	173.32	173.39	173.4	173.41	173.42	173.49	173.5	173.51	173.52	173.59	173.6	173.61	173.62	173.69	173.7	173.71	173.72	173.79	173.8	173.81	173.82	173.89	173.9	173.91	173.92	173.99	174	174.1	174.2	174.3	174.4	174.5	174.6	174.8	174.9	175	175.9	176	176.1	176.2	176.3	176.4	176.5	176.8	176.9	179	180	180.1	180.8	180.9	181	182	182.1	182.8	183	183.2	183.3	183.4	183.5	183.8	183.9	184	184.1	184.2	184.3	184.4	184.8	184.9	185	186	186.9	187	187.1	187.2	187.3	187.4	187.5	187.6	187.7	187.8	187.9	188	188.1	188.2	188.3	188.4	188.5	188.6	188.7	188.8	188.9	189	189.1	189.2	189.3	189.4	189.8	189.9	190	190.1	190.2	190.3	190.4	190.5	190.6	190.7	190.8	190.9	191	191.1	191.2	191.3	191.4	191.5	191.6	191.7	191.8	191.9	192	192.1	192.2	192.3	192.8	192.9	193	194	194.1	194.3	194.4	194.5	194.6	194.8	194.9	195	195.1	195.2	195.3	195.4	195.5	195.8	200	200.01	200.02	200.03	200.04	200.05	200.06	200.07	200.08	200.1	200.11	200.12	200.13	200.14	200.15	200.16	200.17	200.18	200.2	200.21	200.22	200.23	200.24	200.25	200.26	200.27	200.28	200.3	200.31	200.32	200.33	200.34	200.35	200.36	200.37	200.38	200.4	200.41	200.42	200.43	200.44	200.45	200.46	200.47	200.48	200.5	200.51	200.52	200.53	200.54	200.55	200.56	200.57	200.58	200.6	200.61	200.62	200.63	200.64	200.65	200.66	200.67	200.68	200.7	200.71	200.72	200.73	200.74	200.75	200.76	200.77	200.78	200.8	200.81	200.82	200.83	200.84	200.85	200.86	200.87	200.88	201	201.01	201.02	201.03	201.04	201.05	201.06	201.07	201.08	201.1	201.11	201.12	201.13	201.14	201.15	201.16	201.17	201.18	201.2	201.21	201.22	201.23	201.24	201.25	201.26	201.27	201.28	201.4	201.41	201.42	201.43	201.44	201.45	201.46	201.47	201.48	201.5	201.51	201.52	201.53	201.54	201.55	201.56	201.57	201.58	201.6	201.61	201.62	201.63	201.64	201.65	201.66	201.67	201.68	201.7	201.71	201.72	201.73	201.74	201.75	201.76	201.77	201.78	201.9	201.91	201.92	201.93	201.94	201.95	201.96	201.97	201.98	202	202.01	202.02	202.03	202.04	202.05	202.06	202.07	202.08	202.1	202.11	202.12	202.13	202.14	202.15	202.16	202.17	202.18	202.2	202.21	202.22	202.23	202.24	202.25	202.26	202.27	202.28	202.3	202.31	202.32	202.33	202.34	202.35	202.36	202.37	202.38	202.4	202.41	202.42	202.43	202.44	202.45	202.46	202.47	202.48	202.5	202.51	202.52	202.53	202.54	202.55	202.56	202.57	202.58	202.6	202.61	202.62	202.63	202.64	202.65	202.66	202.67	202.68	202.7	202.71	202.72	202.73	202.74	202.75	202.76	202.77	202.78	202.8	202.81	202.82	202.83	202.84	202.85	202.86	202.87	202.88	202.9	202.91	202.92	202.93	202.94	202.95	202.96	202.97	202.98	203	203.01	203.02	203.1	203.11	203.12	203.8	203.81	203.82	204	204.01	204.02	204.1	204.11	204.12	204.2	204.21	204.22	204.8	204.81	204.82	204.9	204.91	204.92	205	205.01	205.02	205.1	205.11	205.12	205.2	205.21	205.22	205.3	205.31	205.32	205.8	205.81	205.82	205.9	205.91	205.92	206	206.01	206.02	206.1	206.11	206.12	206.2	206.21	206.22	206.8	206.81	206.82	206.9	206.91	206.92	207	207.01	207.02	207.1	207.11	207.12	207.2	207.21	207.22	207.8	207.81	207.82	208	208.01	208.02	208.1	208.11	208.12	208.2	208.21	208.22	208.8	208.81	208.82	208.9	208.91	208.92 196	196.1	196.2	196.3	196.5	196.6	196.8	196.9	197	197.1	197.2	197.3	197.4	197.5	197.6	197.7	197.8	198	198.1	198.2	198.3	198.4	198.5	198.6	198.7	198.8	198.81	198.82	198.89	199	199.1

Obesity	278.01	278	278

**Supplementary Table 2. Calculations for measures of variability.**

| **Variability measure** | **Definition** |
| --- | --- |
| Standard deviation |  |
| Coefficient of Variation | $\frac{SD}{individual mean}$ |
| Root mean square |  |

**Supplementary Table 3. Baseline and clinical characteristics comparisons of type-2 diabetes patients with/without adverse events.**

* for p≤ 0.05, ** for p ≤ 0.01, *** for p ≤ 0.001; ACEI: angiotensin-converting-enzyme inhibitors, ARB: angiotensin II receptor blockers, CV: cardiovascular, LDL: low density lipoprotein cholesterol, HDL: high density lipoprotein cholesterol, TC: total cholesterol, TG: triglyceride, NLR: neutrophil-to-lymphocyte, AALP: albumin-to-alkaline phosphatase, SD: standard deviation, SMD: standard mean difference.

| **Characteristics** | **All-cause mortality (N=10182)**  **Median (IQR);N or Count(%)** | **Cardiovascular mortality (N=1966)**  **Median (IQR);N or Count(%)** | **New onset heart failure (N=1171)**  **Median (IQR);N or Count(%)** | **P value** |
| --- | --- | --- | --- | --- |
| ***Demographics*** |  |  |  |  |
| Male gender | 5333.0(52.37%) | 978.0(49.74%) | 546.0(46.62%) | <0.0001*** |
| Female gender | 4849.0(47.62%) | 988.0(50.25%) | 625.0(53.37%) | <0.0001*** |
| Baseline age, years | 77.52(70.05-83.06);n=10182 | 78.0(70.05-83.06);n=1966 | 79.05(71.05-84.06);n=1171 | <0.0001*** |
| <50 | 233.0(2.28%) | 65.0(3.30%) | 24.0(2.04%) | <0.0001*** |
| [50-60] | 702.0(6.89%) | 121.0(6.15%) | 72.0(6.14%) | <0.0001*** |
| [60-70] | 1492.0(14.65%) | 298.0(15.15%) | 154.0(13.15%) | <0.0001*** |
| [70-80] | 3691.0(36.25%) | 691.0(35.14%) | 371.0(31.68%) | <0.0001*** |
| >80 | 4065.0(39.92%) | 791.0(40.23%) | 550.0(46.96%) | <0.0001*** |
| ***Past comorbidities*** |  |  |  |  |
| CHA-DS-VASc Score | 3.0(2.0-4.0);n=10182 | 3.0(2.0-5.0);n=1966 | 5.0(4.0-6.0);n=1171 | 0.2351 |
| Charlson standard comorbidity index | 4.0(3.0-5.0);n=10182 | 4.0(3.0-6.0);n=1966 | 6.0(5.0-8.0);n=1171 | <0.0001*** |
| Number of comorbidities | 2.0(1.0-5.0);n=10182 | 3.0(2.0-5.0);n=1966 | 7.0(5.0-8.0);n=1171 | <0.0001*** |
| Systemic embolism | 22.0(0.21%) | 8.0(0.40%) | 5.0(0.42%) | 0.1178 |
| Hypertension | 5861.0(57.56%) | 1203.0(61.19%) | 844.0(72.07%) | <0.0001*** |
| Coronary heart disease | 2710.0(26.61%) | 764.0(38.86%) | 692.0(59.09%) | <0.0001*** |
| Atrial fibrillation | 694.0(6.81%) | 221.0(11.24%) | 332.0(28.35%) | <0.0001*** |
| Hemiplegia or paraplegia | 282.0(2.76%) | 55.0(2.79%) | 33.0(2.81%) | <0.0001*** |
| Renal diseases | 860.0(8.44%) | 213.0(10.83%) | 216.0(18.44%) | <0.0001*** |
| Osteoporosis | 5.0(0.04%) | 1.0(0.05%) | 1.0(0.08%) | 0.7291 |
| Liver diseases | 79.0(0.77%) | 12.0(0.61%) | 17.0(1.45%) | 0.1638 |
| Ventricular tachycardia/fibrillation | 129.0(1.26%) | 44.0(2.23%) | 61.0(5.20%) | 0.0013** |
| Dementia and Alzheimer | 635.0(6.23%) | 92.0(4.67%) | 56.0(4.78%) | <0.0001*** |
| Anemia | 6322.0(62.08%) | 1235.0(62.81%) | 773.0(66.01%) | <0.0001*** |
| Acute myocardial infarction | 452.0(4.43%) | 146.0(7.42%) | 233.0(19.89%) | <0.0001*** |
| Chronic obstructive pulmonary disease | 504.0(4.94%) | 97.0(4.93%) | 155.0(13.23%) | <0.0001*** |
| Ischemic heart disease | 1648.0(16.18%) | 480.0(24.41%) | 645.0(55.08%) | <0.0001*** |
| Peripheral vascular disease | 153.0(1.50%) | 40.0(2.03%) | 34.0(2.90%) | <0.0001*** |
| Stroke/transient ischemic attack | 1130.0(11.09%) | 222.0(11.29%) | 191.0(16.31%) | <0.0001*** |
| Gastrointestinal bleeding | 536.0(5.26%) | 117.0(5.95%) | 135.0(11.52%) | <0.0001*** |
| Cancer | 82.0(0.80%) | 9.0(0.45%) | 7.0(0.59%) | 0.9553 |
| Obesity | 32.0(0.31%) | 6.0(0.30%) | 10.0(0.85%) | 0.9939 |
| ***Drug prescriptions*** |  |  |  |  |
| Number of diabetes mellitus drugs | 1.0(1.0-2.0);n=10182 | 1.0(1.0-2.0);n=1966 | 1.0(1.0-2.0);n=1171 | <0.0001*** |
| Number of CV drugs | 2.0(1.0-3.0);n=10182 | 3.0(2.0-4.0);n=1966 | 3.0(2.0-4.0);n=1171 | <0.0001*** |
| ACEI/ARB | 5847.0(57.42%) | 1280.0(65.10%) | 947.0(80.87%) | <0.0001*** |
| Beta blockers | 4802.0(47.16%) | 1069.0(54.37%) | 682.0(58.24%) | <0.0001*** |
| Calcium channel blockers | 5718.0(56.15%) | 1169.0(59.46%) | 634.0(54.14%) | <0.0001*** |
| Diuretics | 4643.0(45.60%) | 1052.0(53.50%) | 996.0(85.05%) | <0.0001*** |
| Stains | 3539.0(34.75%) | 851.0(43.28%) | 550.0(46.96%) | 0.4032 |
| Metformin | 1949.0(19.14%) | 403.0(20.49%) | 229.0(19.55%) | <0.0001*** |
| Sulfadiazine | 8784.0(86.26%) | 1733.0(88.14%) | 1119.0(95.55%) | <0.0001*** |
| Insulin | 1421.0(13.95%) | 296.0(15.05%) | 240.0(20.49%) | <0.0001*** |
| Sulphonylurea | 2341.0(22.99%) | 472.0(24.00%) | 290.0(24.76%) | <0.0001*** |
| Gliclazide | 7536.0(74.01%) | 1499.0(76.24%) | 990.0(84.54%) | <0.0001*** |
| Meglitinide | 727.0(7.14%) | 140.0(7.12%) | 115.0(9.82%) | <0.0001*** |
| Alpha-glucosidase inhibitors | 247.0(2.42%) | 60.0(3.05%) | 40.0(3.41%) | 0.0002*** |
| Other anti-diabetic drugs | 9121.0(89.57%) | 1806.0(91.86%) | 1171.0(100.00%) | <0.0001*** |
| ***Subclinical biomarkers*** |  |  |  |  |
| NLR ratio | 3.44(2.3-5.67);n=8968 | 3.4(2.35-5.4);n=1716 | 3.64(2.47-5.87);n=1075 | <0.0001*** |
| AALP ratio | 0.42(0.32-0.54);n=10182 | 0.43(0.33-0.56);n=1966 | 0.41(0.31-0.53);n=1171 | <0.0001*** |
| LDL/HDL ratio | 2.46(1.8-3.25);n=4139 | 2.49(1.84-3.3);n=902 | 2.41(1.78-3.33);n=548 | 0.0348* |
| TC/HDL ratio | 3.93(3.15-4.95);n=7726 | 4.0(3.2-5.0);n=1615 | 3.82(3.06-4.72);n=1026 | 0.0862 |
| Triglyceride/HDL | 1.22(0.76-1.97);n=7726 | 1.27(0.79-2.01);n=1615 | 1.2(0.75-1.84);n=1026 | 0.9993 |
| ***Complete blood counts*** |  |  |  |  |
| Lymphocyte, x10^9/L | 1.5(1.07-1.96);n=8968 | 1.5(1.1-1.98);n=1716 | 1.4(1.0-1.88);n=1075 | <0.0001*** |
| Neutrophil, x10^9/L | 5.2(3.9-7.1);n=8968 | 5.2(3.96-6.9);n=1716 | 5.14(3.9-6.9);n=1075 | <0.0001*** |
| Potassium, mmol/L | 4.2(3.8-4.6);n=10162 | 4.2(3.8-4.6);n=1966 | 4.2(3.8-4.59);n=1170 | 0.5185 |
| Albumin, g/L | 37.0(32.0-40.7);n=10182 | 37.0(33.0-41.0);n=1966 | 37.0(33.0-40.8);n=1171 | <0.0001*** |
| Sodium, mmol/L | 139.02(137.0-141.53);n=10162 | 140.0(137.0-142.0);n=1966 | 140.0(137.0-142.0);n=1170 | <0.0001*** |
| Urea, mmol/L | 7.82(5.54-12.0);n=10136 | 8.6(6.0-13.1);n=1960 | 9.32(6.55-14.25);n=1164 | <0.0001*** |
| Creatinine, umol/L | 113.0(82.0-168.0);n=10164 | 124.0(90.0-175.0);n=1966 | 128.0(96.5-182.0);n=1170 | <0.0001*** |
| Total protein, g/L | 72.31(67.0-77.69);n=10176 | 72.08(67.0-77.45);n=1966 | 72.0(66.8-77.05);n=1171 | <0.0001*** |
| Alkaline phosphatase, U/L | 85.0(69.0-108.0);n=9934 | 84.0(68.0-106.0);n=1916 | 87.0(70.0-110.0);n=1155 | <0.0001*** |
| Aspartate transaminase, U/L | 24.0(18.0-33.0);n=3679 | 24.0(18.0-33.0);n=697 | 25.0(18.0-34.0);n=449 | <0.0001*** |
| Alanine transamise, U/L | 18.0(13.0-27.0);n=8822 | 17.0(13.0-25.0);n=1701 | 17.0(12.0-26.0);n=1060 | <0.0001*** |
| Bilirubin, umol/L | 10.3(7.0-15.0);n=10175 | 10.68(7.0-15.0);n=1966 | 11.0(7.14-16.0);n=1171 | <0.0001*** |
| ***Lipid/glycemic profile and variations*** |  |  |  |  |
| Fast glucose, mmol/L | 7.72(6.2-10.5);n=9945 | 7.8(6.1-10.7);n=1929 | 8.0(6.2-10.75);n=1166 | 0.0008*** |
| SD of fast glucose | 1.48(0.91-2.32);n=6701 | 1.5(0.94-2.34);n=1399 | 1.62(1.06-2.49);n=914 | <0.0001*** |
| HbA1c, g/dL | 11.6(10.1-13.1);n=9692 | 11.5(10.1-13.0);n=1874 | 11.25(9.9-12.9);n=1136 | <0.0001*** |
| SD of HbA1c | 0.75(0.46-1.22);n=7134 | 0.76(0.47-1.25);n=1464 | 0.78(0.49-1.25);n=968 | <0.0001*** |
| Low-density lipoprotein, mmol/L | 2.71(2.14-3.36);n=4651 | 2.7(2.14-3.4);n=988 | 2.65(2.14-3.31);n=593 | <0.0001*** |
| SD of LDL | 0.57(0.34-0.91);n=1743 | 0.55(0.3-0.93);n=412 | 0.65(0.37-0.96);n=224 | <0.0001*** |
| High-density lipoprotein, mmol/L | 1.11(0.91-1.38);n=7726 | 1.08(0.9-1.36);n=1615 | 1.1(0.9-1.34);n=1026 | <0.0001*** |
| SD of HDL | 0.15(0.1-0.22);n=4623 | 0.15(0.1-0.21);n=1047 | 0.16(0.11-0.23);n=675 | <0.0001*** |
| Total cholesterol, mmol/L | 4.47(3.79-5.24);n=8576 | 4.42(3.74-5.2);n=1750 | 4.24(3.6-5.04);n=1098 | <0.0001*** |
| SD of TC | 0.65(0.42-0.98);n=5588 | 0.66(0.44-0.98);n=1237 | 0.68(0.45-0.96);n=823 | <0.0001*** |
| Triglyceride, mmol/L | 1.34(0.96-1.92);n=8546 | 1.37(0.98-1.94);n=1746 | 1.27(0.92-1.86);n=1096 | 0.0027** |
| SD of TG | 0.41(0.24-0.72);n=5549 | 0.43(0.26-0.73);n=1229 | 0.43(0.26-0.72);n=820 | 0.3114 |
| ***Alkaline phosphatase variability measures*** |  |  |  |  |
| Baseline, U/L | 85.0(69.0-108.0);n=10182 | 84.0(68.0-106.0);n=1966 | 88.0(70.0-111.0);n=1171 | <0.0001*** |
| Low tertile | 3006.0(29.52%) | 595.0(30.26%) | 318.0(27.15%) | <0.0001*** |
| Middle tertile | 3485.0(34.22%) | 694.0(35.30%) | 394.0(33.64%) | 0.0010*** |
| High tertile | 3691.0(36.25%) | 677.0(34.43%) | 459.0(39.19%) | <0.0001*** |
| Latest, U/L | 85.0(69.0-108.0);n=10182 | 84.0(68.0-106.0);n=1966 | 87.0(70.0-110.0);n=1171 | <0.0001*** |
| Maximum, U/L | 100.0(79.0-133.0);n=10182 | 98.5(78.5-131.0);n=1966 | 105.0(83.0-139.5);n=1171 | <0.0001*** |
| Minimal, U/L | 72.0(59.0-89.0);n=10182 | 71.0(58.0-88.0);n=1966 | 72.0(59.0-89.0);n=1171 | <0.0001*** |
| Mean, U/L | 86.0(70.33-109.0);n=10182 | 85.54(69.83-107.04);n=1966 | 88.33(72.0-110.68);n=1171 | <0.0001*** |
| Median, U/L | 85.0(69.5-107.0);n=10182 | 84.0(69.0-105.25);n=1966 | 87.5(71.0-108.75);n=1171 | <0.0001*** |
| Variance | 110.33(32.33-338.0);n=10182 | 109.71(32.7-319.93);n=1966 | 128.25(42.33-414.51);n=1171 | <0.0001*** |
| SD | 10.5(5.69-18.38);n=10182 | 10.47(5.72-17.89);n=1966 | 11.32(6.51-20.36);n=1171 | <0.0001*** |
| RMS | 86.73(70.9-110.42);n=10182 | 86.17(70.28-108.39);n=1966 | 88.97(72.73-113.1);n=1171 | <0.0001*** |
| CV | 0.11(0.06-0.17);n=10182 | 0.11(0.06-0.17);n=1966 | 0.12(0.07-0.18);n=1171 | <0.0001*** |

**Supplementary Table 4. Univariable Cox regression models for new onset heart failure, cardiovascular mortality, and all-cause mortality.**

* for p≤ 0.05, ** for p ≤ 0.01, *** for p ≤ 0.001; ACEI: angiotensin-converting-enzyme inhibitors, ARB: angiotensin II receptor blockers, CV: cardiovascular, LDL: low density lipoprotein cholesterol, HDL: high density lipoprotein cholesterol, TC: total cholesterol, TG: triglyceride, NLR: neutrophil-to-lymphocyte, AALP: albumin-to-alkaline phosphatase, SD: standard deviation, SMD: standard mean difference.

| **Characteristics** | **All-cause mortality**  **HR [95% CI];P value** | **Cardiovascular mortality**  **HR [95% CI];P value** | **New onset heart failure**  **HR [95% CI];P value** |
| --- | --- | --- | --- |
| ***Demographics*** |  |  |  |
| Male gender | 1.0[Reference] | 1.0[Reference] | 1.0[Reference] |
| Female gender | 1.02[0.98-1.06];0.4388 | 1.13[1.03-1.23];0.0078** | 1.28[1.14-1.44];<0.0001*** |
| Baseline age, years | 1.05[1.05-1.06];<0.0001*** | 1.05[1.04-1.05];<0.0001*** | 1.05[1.05-1.06];<0.0001*** |
| <50 | 1.0[Reference] | 1.0[Reference] | 1.0[Reference] |
| [50-60] | 0.37[0.35-0.40];<0.0001*** | 0.34[0.28-0.41];<0.0001*** | 0.37[0.29-0.47];<0.0001*** |
| [60-70] | 0.61[0.57-0.64];<0.0001*** | 0.64[0.57-0.72];<0.0001*** | 0.57[0.48-0.68];<0.0001*** |
| [70-80] | 1.14[1.10-1.19];<0.0001*** | 1.08[0.99-1.19];0.0963 | 0.93[0.82-1.05];0.2312 |
| >80 | 2.46[2.36-2.56];<0.0001*** | 2.40[2.19-2.64];<0.0001*** | 2.73[2.43-3.07];<0.0001*** |
| ***Past comorbidities*** |  |  |  |
| CHA-DS-VASc Score | 1.26[1.24-1.27];<0.0001*** | 1.33[1.30-1.36];<0.0001*** | 2.04[1.98-2.10];<0.0001*** |
| Charlson standard comorbidity index | 1.26[1.25-1.27];<0.0001*** | 1.31[1.28-1.33];<0.0001*** | 1.72[1.69-1.76];<0.0001*** |
| Number of comorbidities | 1.15[1.14-1.16];<0.0001*** | 1.24[1.22-1.26];<0.0001*** | 1.71[1.67-1.74];<0.0001*** |
| Systemic embolism | 1.39[0.92-2.11];0.1225 | 2.58[1.29-5.16];0.0075** | 2.77[1.15-6.68];0.0229* |
| Hypertension | 1.65[1.58-1.71];<0.0001*** | 1.89[1.72-2.07];<0.0001*** | 2.95[2.59-3.35];<0.0001*** |
| Coronary heart disease | 1.27[1.21-1.33];<0.0001*** | 2.21[2.02-2.42];<0.0001*** | 5.06[4.50-5.69];<0.0001*** |
| Atrial fibrillation | 1.63[1.51-1.77];<0.0001*** | 2.78[2.42-3.20];<0.0001*** | 9.03[7.94-10.26];<0.0001*** |
| Hemiplegia or paraplegia | 1.51[1.34-1.70];<0.0001*** | 1.50[1.15-1.96];0.0029** | 1.41[1.00-1.99];0.0525 |
| Renal diseases | 1.95[1.81-2.09];<0.0001*** | 2.50[2.17-2.89];<0.0001*** | 4.21[3.63-4.88];<0.0001*** |
| Osteoporosis | 1.53[0.64-3.67];0.3424 | 1.53[0.22-10.89];0.6695 | 2.41[0.34-17.12];0.3795 |
| Liver diseases | 1.23[0.98-1.53];0.0684 | 0.96[0.54-1.69];0.8838 | 2.19[1.36-3.54];0.0013** |
| Ventricular tachycardia/fibrillation | 1.30[1.10-1.55];0.0028** | 2.30[1.71-3.10];<0.0001*** | 5.72[4.42-7.40];<0.0001*** |
| Dementia and Alzheimer | 2.44[2.25-2.65];<0.0001*** | 1.73[1.40-2.13];<0.0001*** | 1.41[1.08-1.85];0.0125* |
| Anemia | 2.40[2.31-2.50];<0.0001*** | 2.42[2.20-2.65];<0.0001*** | 2.46[2.17-2.78];<0.0001*** |
| Acute myocardial infarction | 1.23[1.12-1.35];<0.0001*** | 2.11[1.78-2.49];<0.0001*** | 7.01[6.07-8.09];<0.0001*** |
| Chronic obstructive pulmonary disease | 1.57[1.44-1.72];<0.0001*** | 1.54[1.25-1.89];<0.0001*** | 4.47[3.77-5.29];<0.0001*** |
| Ischemic heart disease | 1.20[1.14-1.26];<0.0001*** | 1.99[1.80-2.21];<0.0001*** | 7.97[7.10-8.94];<0.0001*** |
| Peripheral vascular disease | 2.08[1.77-2.44];<0.0001*** | 2.74[2.01-3.75];<0.0001*** | 3.31[2.35-4.65];<0.0001*** |
| Stroke/transient ischemic attack | 1.44[1.35-1.53];<0.0001*** | 1.45[1.26-1.66];<0.0001*** | 2.12[1.81-2.47];<0.0001*** |
| Gastrointestinal bleeding | 1.34[1.23-1.47];<0.0001*** | 1.51[1.26-1.83];<0.0001*** | 3.06[2.56-3.66];<0.0001*** |
| Cancer | 0.96[0.77-1.20];0.7277 | 0.54[0.28-1.05];0.0692 | 0.71[0.34-1.49];0.3674 |
| Obesity | 0.87[0.61-1.22];0.4156 | 0.84[0.38-1.87];0.6691 | 2.61[1.40-4.86];0.0025** |
| ***Drug prescriptions*** |  |  |  |
| Number of diabetes mellitus drugs | 1.20[1.17-1.24];<0.0001*** | 1.33[1.25-1.41];<0.0001*** | 1.58[1.47-1.71];<0.0001*** |
| Number of CV drugs | 1.11[1.10-1.13];<0.0001*** | 1.33[1.29-1.38];<0.0001*** | 1.78[1.70-1.86];<0.0001*** |
| ACEI/ARB | 1.11[1.06-1.15];<0.0001*** | 1.52[1.39-1.67];<0.0001*** | 3.51[3.03-4.06];<0.0001*** |
| Beta blockers | 1.06[1.02-1.11];0.0020** | 1.42[1.30-1.55];<0.0001*** | 1.66[1.48-1.87];<0.0001*** |
| Calcium channel blockers | 1.39[1.34-1.44];<0.0001*** | 1.57[1.44-1.72];<0.0001*** | 1.21[1.08-1.36];0.0010** |
| Diuretics | 1.77[1.70-1.84];<0.0001*** | 2.40[2.19-2.62];<0.0001*** | 11.55[9.83-13.58];<0.0001*** |
| Stains | 0.81[0.78-0.85];<0.0001*** | 1.17[1.07-1.28];0.0005*** | 1.43[1.27-1.60];<0.0001*** |
| Metformin | 1.25[1.19-1.31];<0.0001*** | 1.35[1.21-1.51];<0.0001*** | 1.24[1.08-1.44];0.0032** |
| Sulfadiazine | 1.07[1.01-1.14];0.0133* | 1.27[1.10-1.45];0.0007*** | 3.66[2.77-4.84];<0.0001*** |
| Insulin | 1.71[1.61-1.80];<0.0001*** | 1.84[1.62-2.08];<0.0001*** | 2.41[2.09-2.78];<0.0001*** |
| Sulphonylurea | 1.27[1.21-1.33];<0.0001*** | 1.33[1.20-1.48];<0.0001*** | 1.33[1.16-1.52];<0.0001*** |
| Gliclazide | 1.09[1.05-1.14];0.0001*** | 1.23[1.11-1.36];0.0001*** | 2.08[1.78-2.44];<0.0001*** |
| Meglitinide | 1.38[1.28-1.48];<0.0001*** | 1.36[1.14-1.61];0.0005*** | 1.81[1.49-2.19];<0.0001*** |
| Alpha-glucosidase inhibitors | 1.25[1.10-1.42];0.0005*** | 1.57[1.21-2.03];0.0006*** | 1.70[1.24-2.32];0.0010** |
| Other anti-diabetic drugs | 1.23[1.15-1.31];<0.0001*** | 1.60[1.36-1.88];<0.0001*** | - |
| ***Subclinical biomarkers*** |  |  |  |
| NLR ratio | 1.021[1.019-1.023];<0.0001*** | 1.02[1.01-1.02];<0.0001*** | 1.02[1.01-1.03];<0.0001*** |
| AALP ratio | 0.15[0.13-0.17];<0.0001*** | 0.23[0.18-0.30];<0.0001*** | 0.18[0.13-0.25];<0.0001*** |
| LDL/HDL ratio | 1.05[1.03-1.07];<0.0001*** | 1.08[1.04-1.13];0.0002*** | 1.06[1.01-1.13];0.0327* |
| TC/HDL ratio | 1.00[0.99-1.02];0.5385 | 1.01[0.99-1.04];0.3196 | 0.94[0.90-0.98];0.0072** |
| Triglyceride/HDL | 1.00[1.00-1.01];0.4874 | 1.00[0.99-1.01];0.9848 | 0.98[0.94-1.02];0.3843 |
| ***Complete blood counts*** |  |  |  |
| Lymphocyte, x10^9/L | 0.72[0.70-0.74];<0.0001*** | 0.75[0.70-0.81];<0.0001*** | 0.63[0.58-0.70];<0.0001*** |
| Neutrophil, x10^9/L | 1.05[1.04-1.05];<0.0001*** | 1.04[1.03-1.06];<0.0001*** | 1.03[1.01-1.05];0.0026** |
| Potassium, mmol/L | 0.99[0.95-1.02];0.4467 | 1.03[0.95-1.12];0.4707 | 0.96[0.86-1.08];0.5070 |
| Albumin, g/L | 0.92[0.92-0.93];<0.0001*** | 0.94[0.93-0.94];<0.0001*** | 0.95[0.94-0.96];<0.0001*** |
| Sodium, mmol/L | 0.95[0.94-0.96];<0.0001*** | 0.97[0.96-0.98];<0.0001*** | 0.99[0.97-1.00];0.0772 |
| Urea, mmol/L | 1.05[1.05-1.06];<0.0001*** | 1.07[1.06-1.07];<0.0001*** | 1.06[1.05-1.07];<0.0001*** |
| Creatinine, umol/L | 1.001[1.001-1.001];<0.0001*** | 1.002[1.001-1.002];<0.0001*** | 1.001[1.001-1.001];<0.0001*** |
| Total protein, g/L | 0.96[0.96-0.97];<0.0001*** | 0.97[0.96-0.97];<0.0001*** | 0.97[0.96-0.98];<0.0001*** |
| Alkaline phosphatase, U/L | 1.002[1.002-1.002];<0.0001*** | 1.001[1.001-1.002];<0.0001*** | 1.001[1.001-1.002];<0.0001*** |
| Aspartate transaminase, U/L | 1.000[1.000-1.000];0.5643 | 1.000[0.999-1.001];0.4711 | 1.000[0.999-1.001];0.8857 |
| Alanine transamise, U/L | 0.99[0.99-1.00];<0.0001*** | 0.98[0.98-0.99];<0.0001*** | 0.99[0.98-0.99];<0.0001*** |
| Bilirubin, umol/L | 1.00[0.99-1.00];0.0001*** | 0.99[0.99-1.00];0.0009*** | 1.00[0.99-1.00];0.3062 |
| ***Lipid/glycemic profile and variations*** |  |  |  |
| Fast glucose, mmol/L | 1.01[1.01-1.02];<0.0001*** | 1.01[1.00-1.02];0.0140* | 1.02[1.01-1.03];0.0048** |
| SD of fast glucose | 1.14[1.12-1.16];<0.0001*** | 1.15[1.11-1.19];<0.0001*** | 1.20[1.16-1.25];<0.0001*** |
| HbA1c, g/dL | 0.80[0.80-0.81];<0.0001*** | 0.80[0.78-0.82];<0.0001*** | 0.81[0.79-0.84];<0.0001*** |
| SD of HbA1c | 1.17[1.14-1.21];<0.0001*** | 1.19[1.12-1.26];<0.0001*** | 1.18[1.09-1.27];<0.0001*** |
| Low-density lipoprotein, mmol/L | 0.96[0.93-0.99];0.0036** | 0.96[0.90-1.02];0.1955 | 0.92[0.85-1.01];0.0720 |
| SD of LDL | 1.36[1.22-1.52];<0.0001*** | 1.24[0.98-1.55];0.0722 | 1.61[1.21-2.14];0.0010** |
| High-density lipoprotein, mmol/L | 0.87[0.82-0.93];<0.0001*** | 0.73[0.64-0.84];<0.0001*** | 0.77[0.65-0.91];0.0030** |
| SD of HDL | 4.33[3.41-5.51];<0.0001*** | 3.07[1.82-5.20];<0.0001*** | 5.29[2.93-9.56];<0.0001*** |
| Total cholesterol, mmol/L | 0.93[0.91-0.95];<0.0001*** | 0.91[0.87-0.95];<0.0001*** | 0.77[0.73-0.82];<0.0001*** |
| SD of TC | 1.25[1.19-1.32];<0.0001*** | 1.25[1.12-1.39];0.0001*** | 1.25[1.09-1.42];0.0009*** |
| Triglyceride, mmol/L | 0.97[0.96-0.99];0.0032** | 0.98[0.94-1.02];0.2357 | 0.92[0.87-0.97];0.0035** |
| SD of TG | 0.99[0.96-1.02];0.5493 | 0.99[0.94-1.05];0.7838 | 0.97[0.90-1.05];0.4628 |
| ***Alkaline phosphatase variability measures*** |  |  |  |
| Baseline, U/L | 1.002[1.002-1.002];<0.0001*** | 1.002[1.001-1.002];<0.0001*** | 1.002[1.001-1.003];<0.0001*** |
| Baseline (High v.s. low or middle tertile) | 1.34[1.29-1.40];<0.0001*** | 1.23[1.12-1.35];<0.0001*** | 1.44[1.28-1.62];<0.0001*** |
| Latest, U/L | 1.002[1.002-1.002];<0.0001*** | 1.001[1.001-1.002];<0.0001*** | 1.001[1.001-1.002];<0.0001*** |
| Maximum, U/L | 1.002[1.002-1.002];<0.0001*** | 1.002[1.001-1.002];<0.0001*** | 1.001[1.001-1.002];<0.0001*** |
| Minimal, U/L | 1.002[1.002-1.003];<0.0001*** | 1.001[1.000-1.003];0.0243* | 1.002[1.000-1.003];0.0232* |
| Mean, U/L | 1.003[1.003-1.003];<0.0001*** | 1.002[1.002-1.003];<0.0001*** | 1.002[1.001-1.003];<0.0001*** |
| Median, U/L | 1.003[1.003-1.003];<0.0001*** | 1.002[1.002-1.003];<0.0001*** | 1.002[1.001-1.003];<0.0001*** |
| Variance | 1.000[1.000-1.000];<0.0001*** | 1.000[1.000-1.000];0.1984 | 1.000[1.000-1.000];0.9126 |
| SD | 1.00[1.00-1.01];<0.0001*** | 1.00[1.00-1.01];<0.0001*** | 1.004[1.002-1.005];<0.0001*** |
| RMS | 1.003[1.003-1.003];<0.0001*** | 1.002[1.002-1.003];<0.0001*** | 1.002[1.001-1.003];<0.0001*** |
| CV | 8.14[7.04-9.42];<0.0001*** | 7.19[5.13-10.06];<0.0001*** | 7.56[5.02-11.37];<0.0001*** |

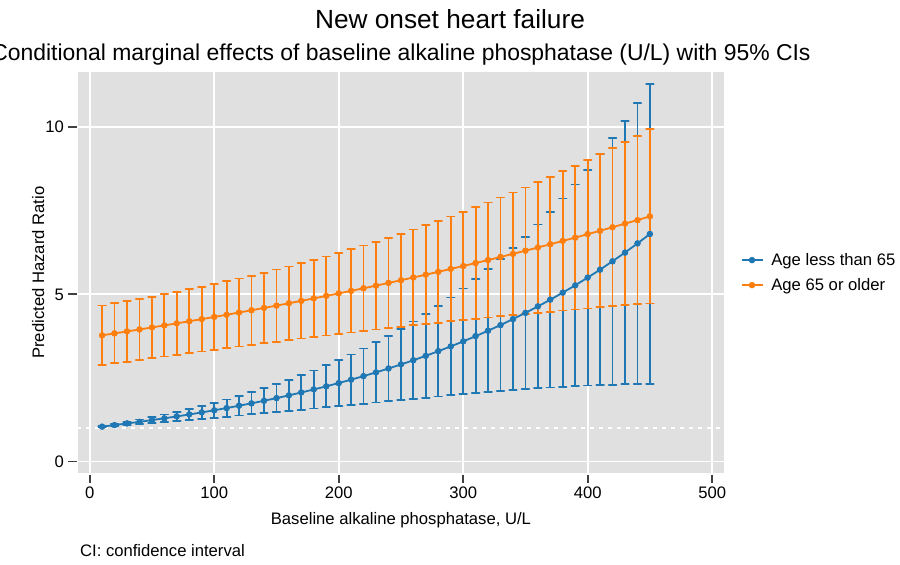

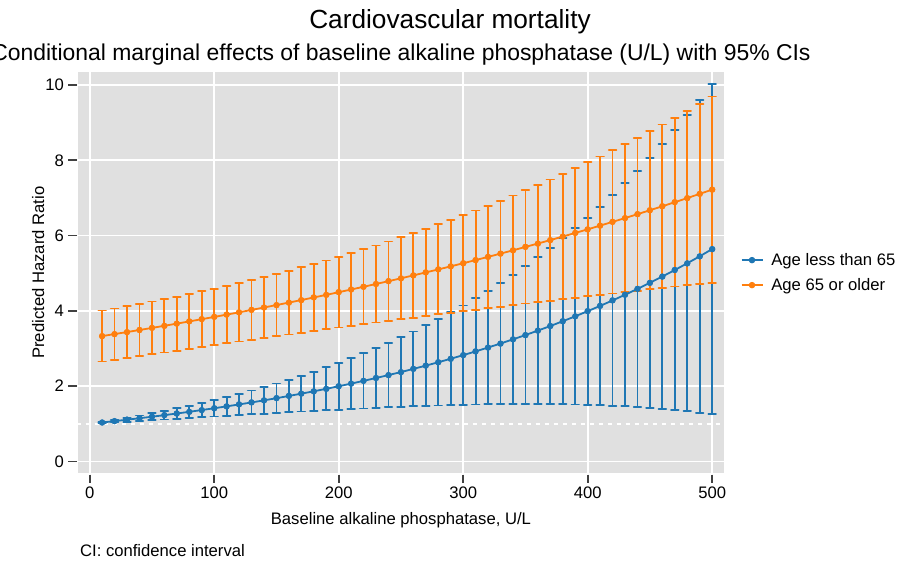

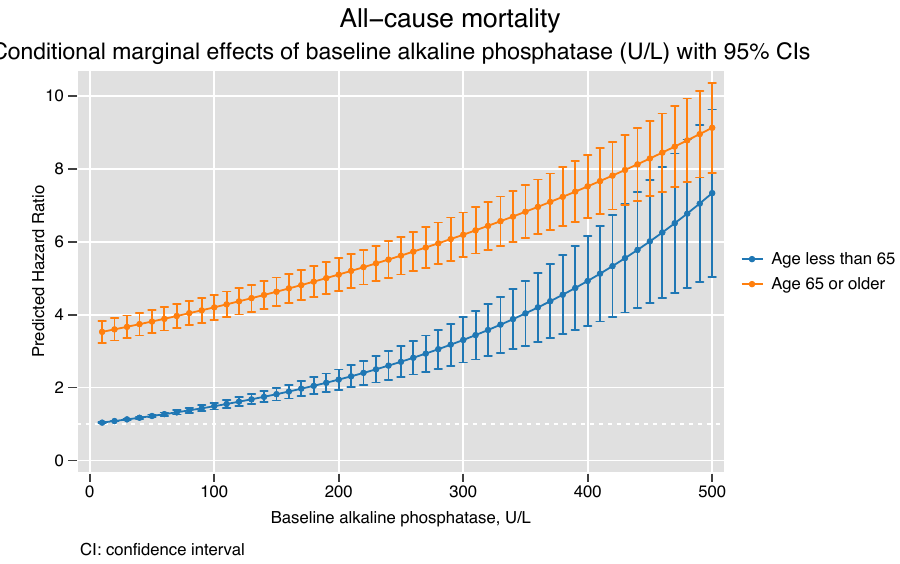

**Supplementary Figure 1A. Marginal effects of baseline alkaline phosphatase to predict new onset heart failure, cardiovascular mortality, and all-cause mortality.**

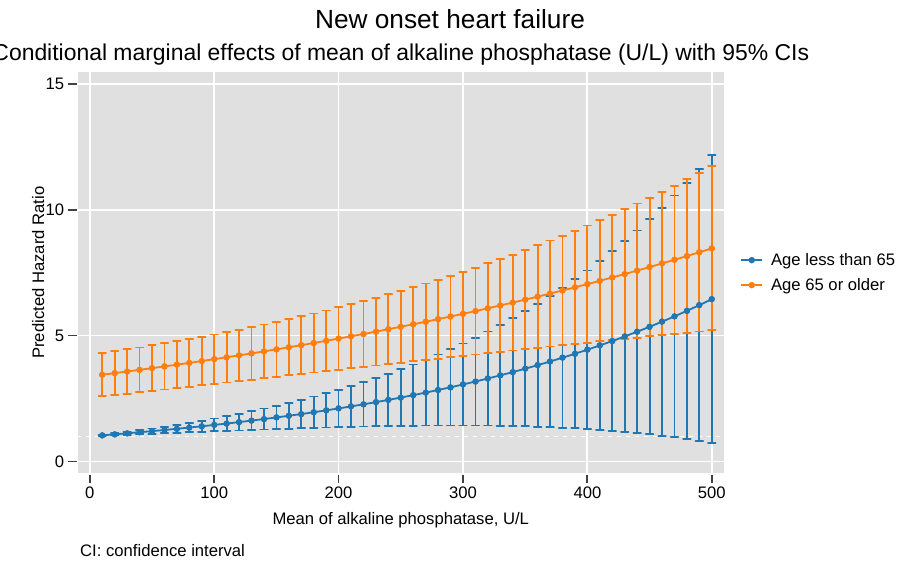

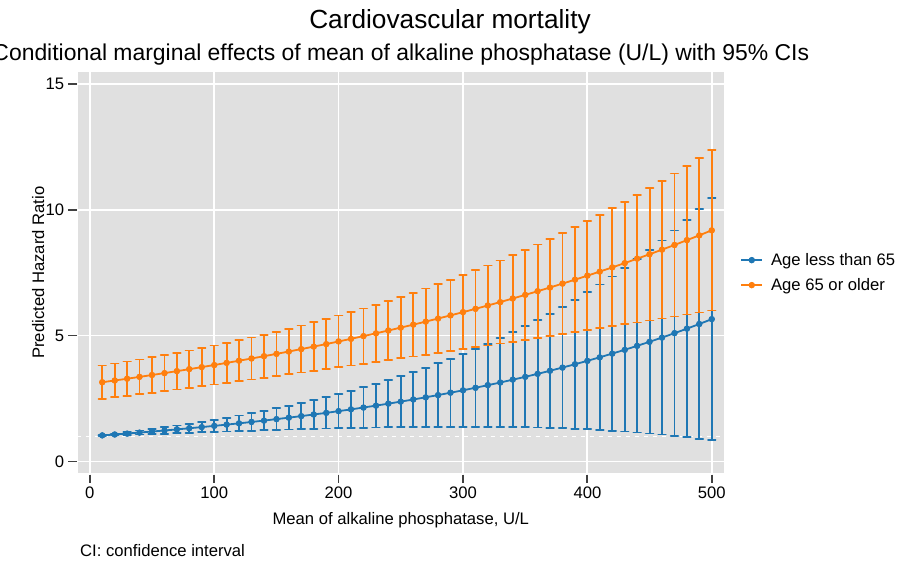

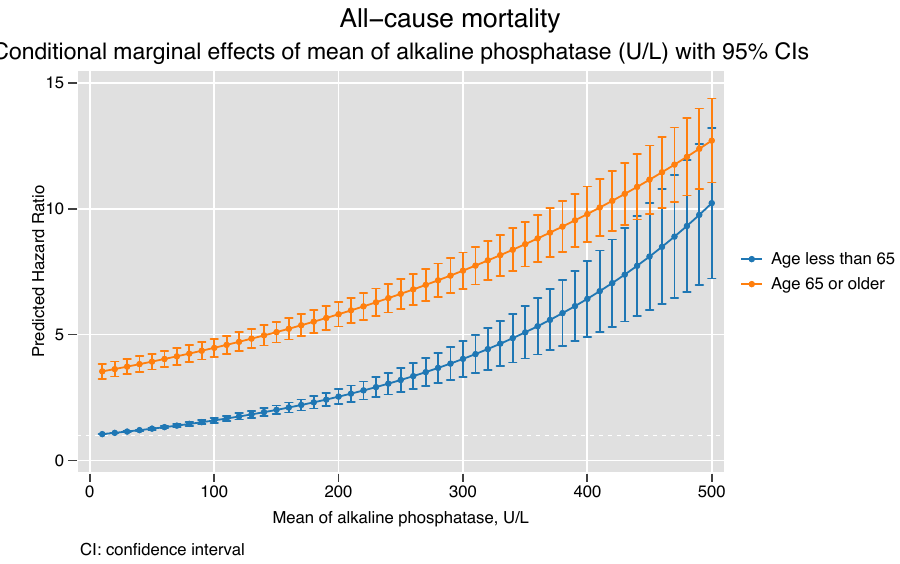

**Supplementary Figure 1B. Marginal effects of mean alkaline phosphatase stratified by patient age to predict new onset heart failure, cardiovascular mortality, and all-cause mortality.**

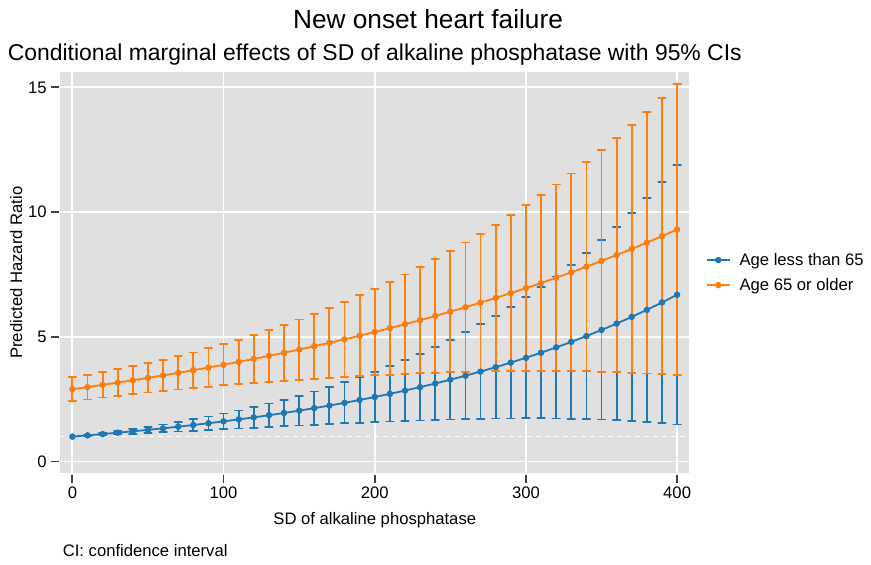

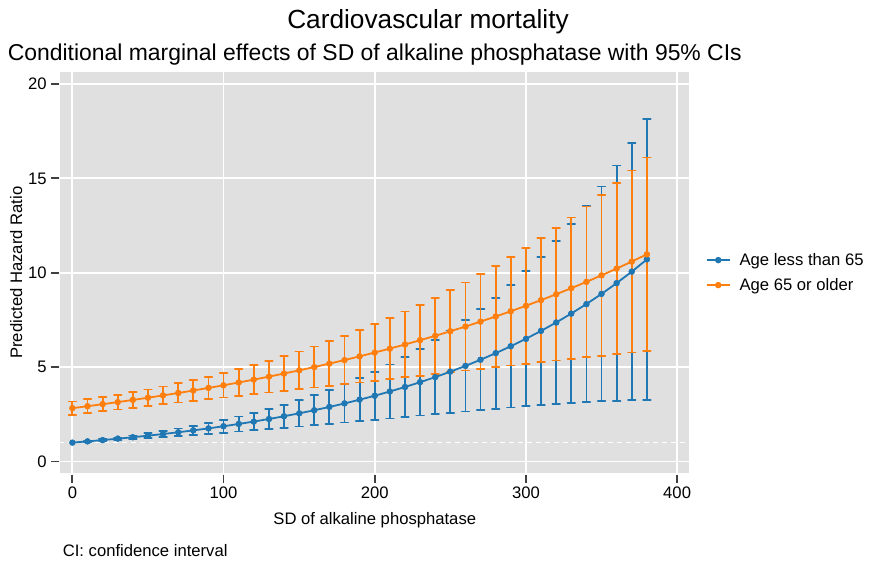

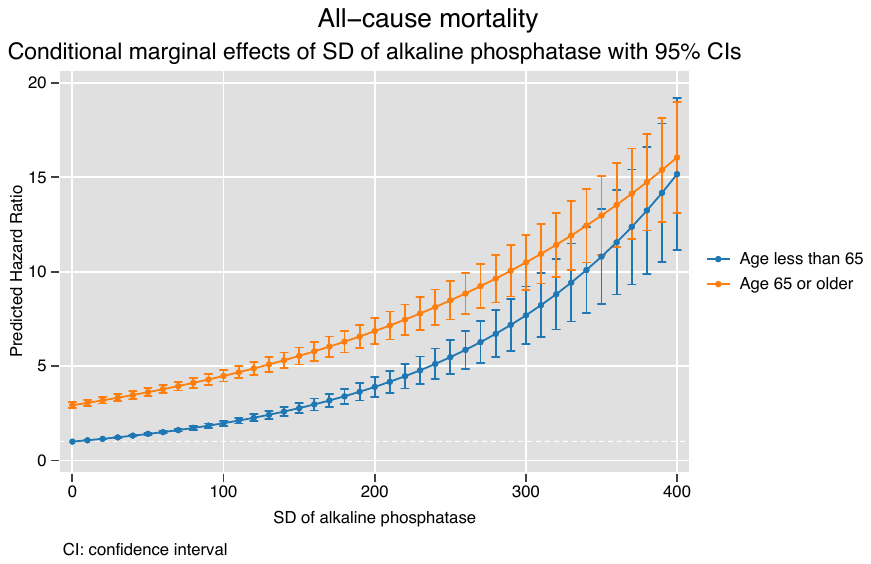

**Supplementary Figure 1C. Marginal effects of SD of alkaline phosphatase stratified by patient age to predict new onset heart failure, cardiovascular mortality, and all-cause mortality.**

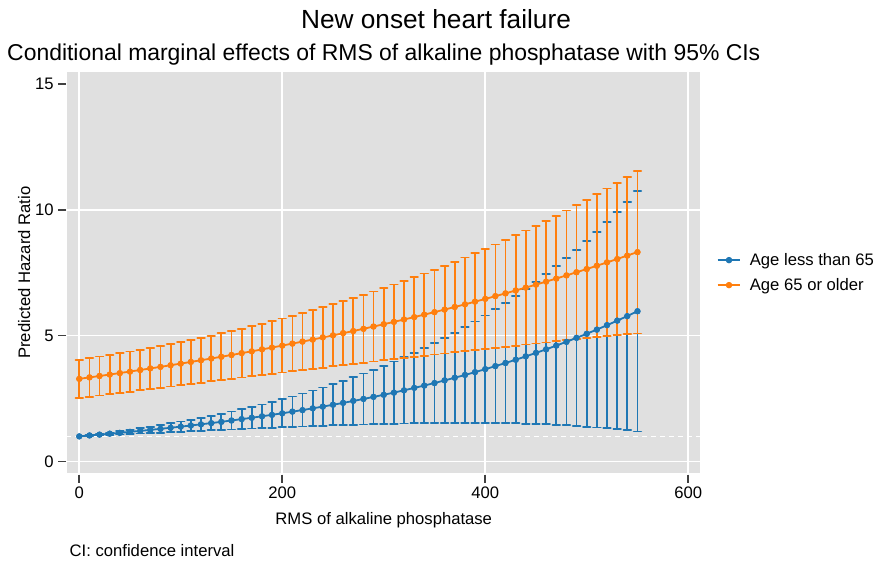

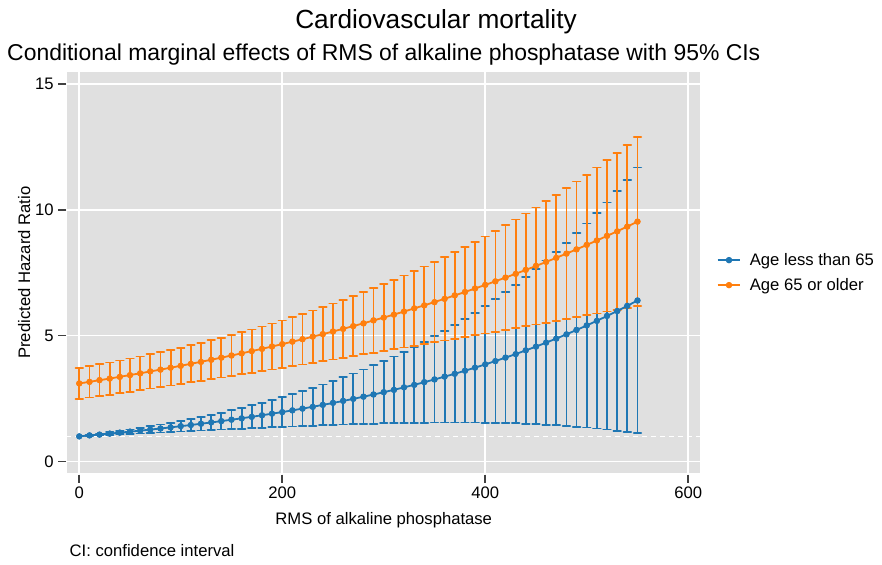

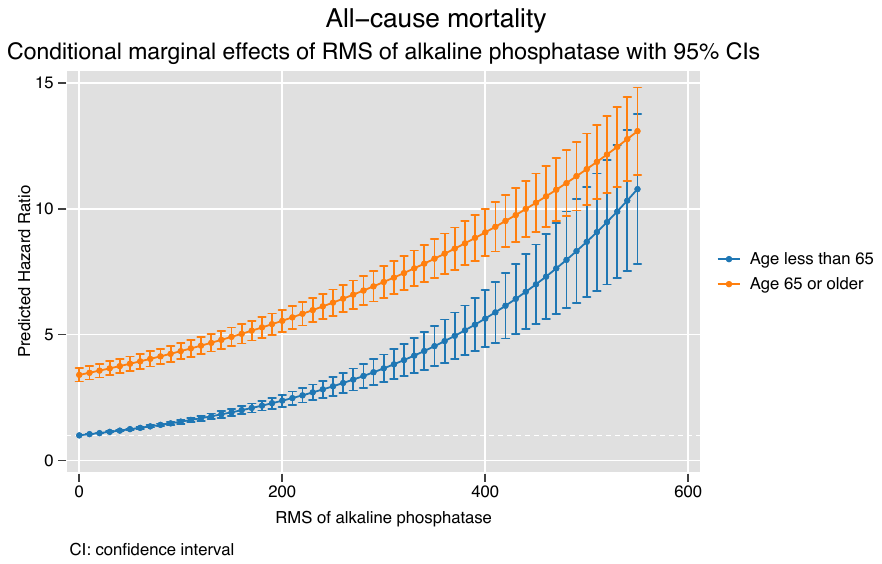

**Supplementary Figure 1D. Marginal effects of RMS of alkaline phosphatase stratified by patient age to predict new onset heart failure, cardiovascular mortality, and all-cause mortality.**

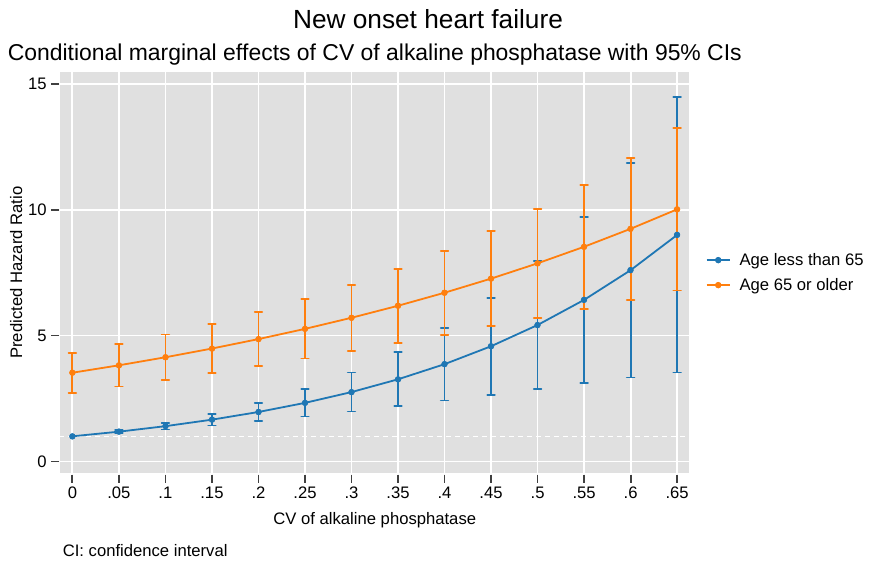

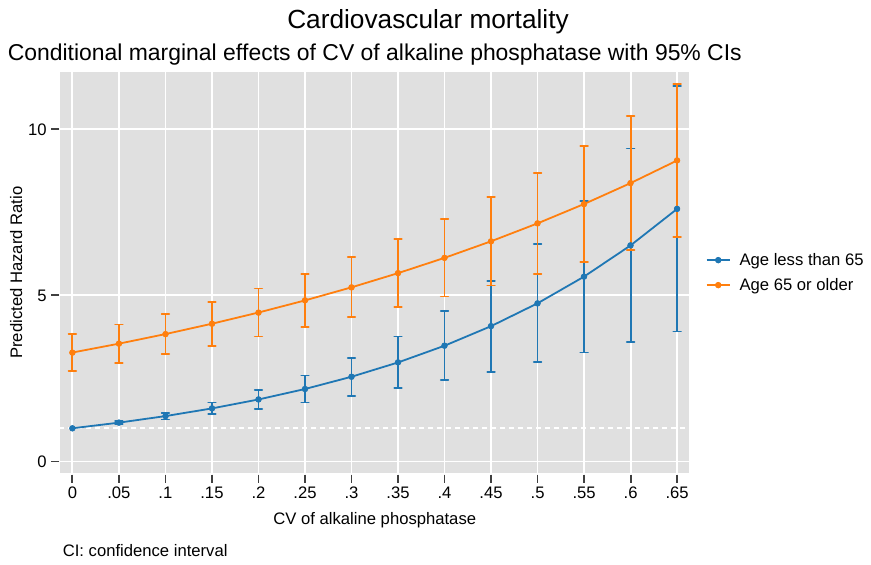

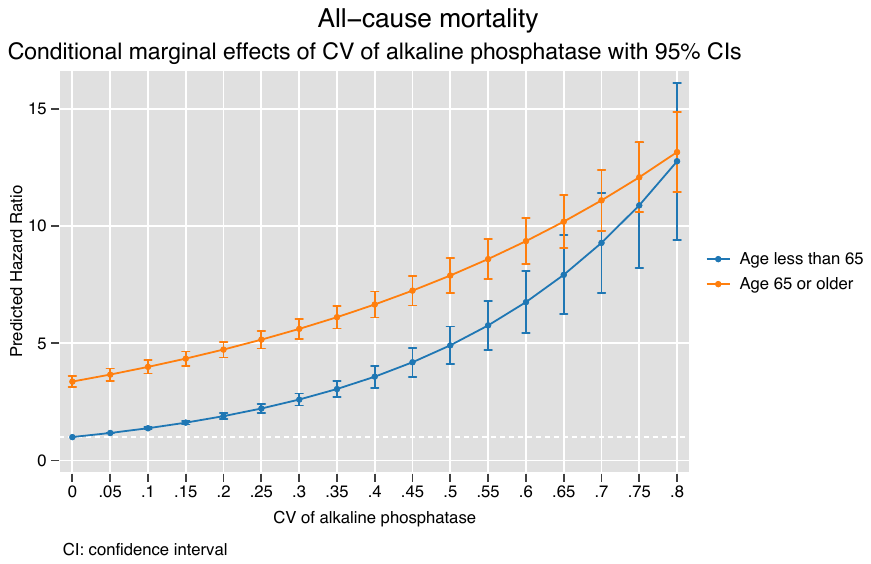

**Supplementary Figure 1E. Marginal effects of CV of alkaline phosphatase stratified by patient age to predict new onset heart failure, cardiovascular mortality, and all-cause mortality.**

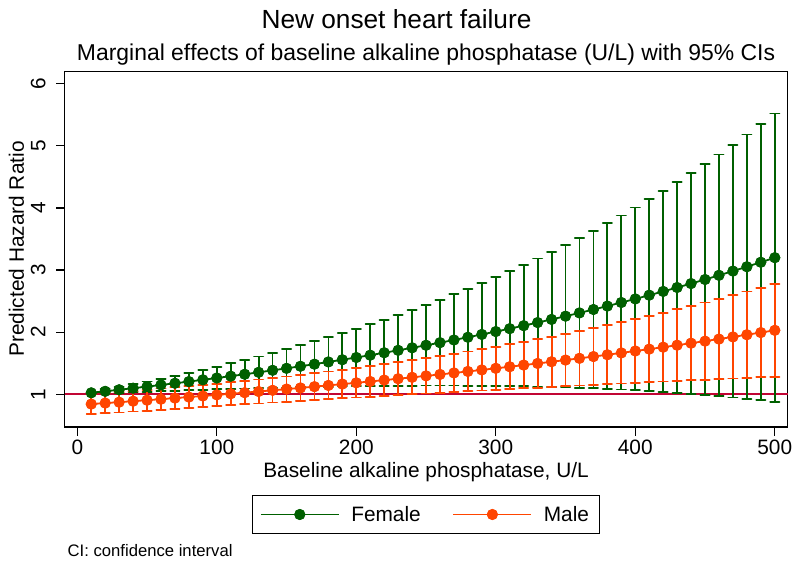

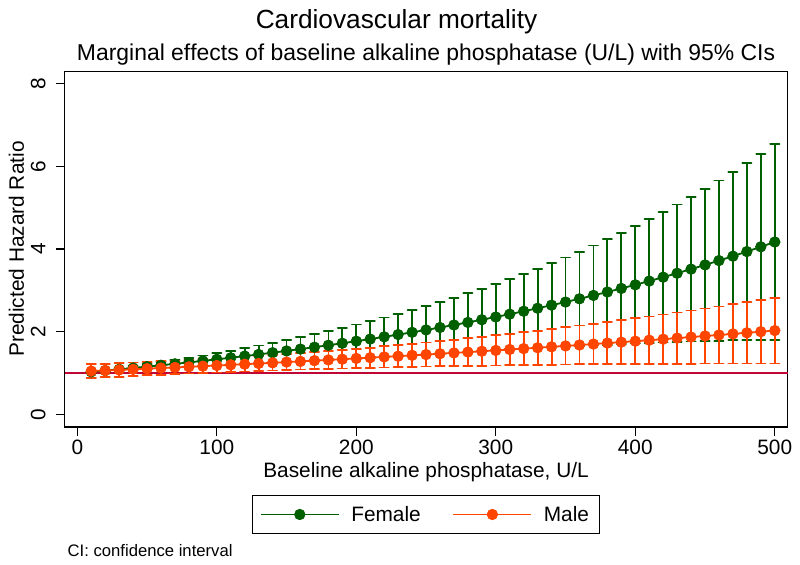

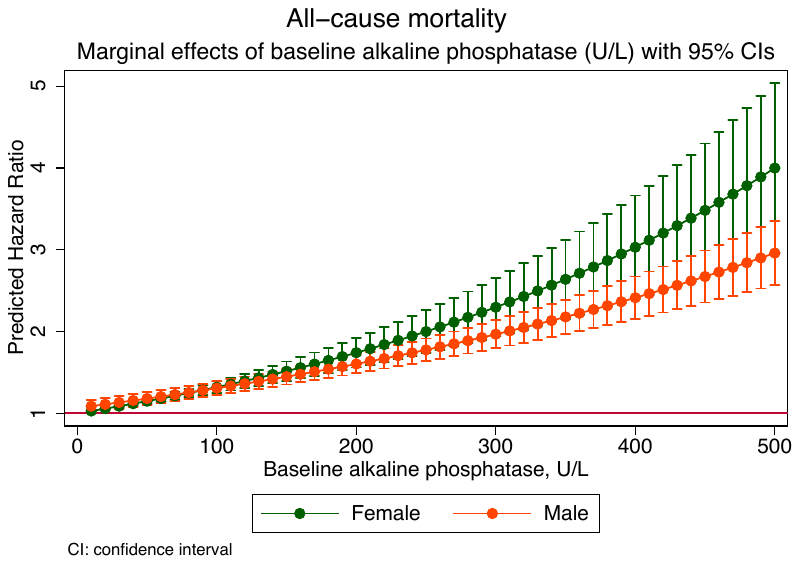

**Supplementary Figure 2A. Marginal effects of baseline alkaline phosphatase stratified by gender to predict new onset heart failure, cardiovascular mortality, and all-cause mortality.**

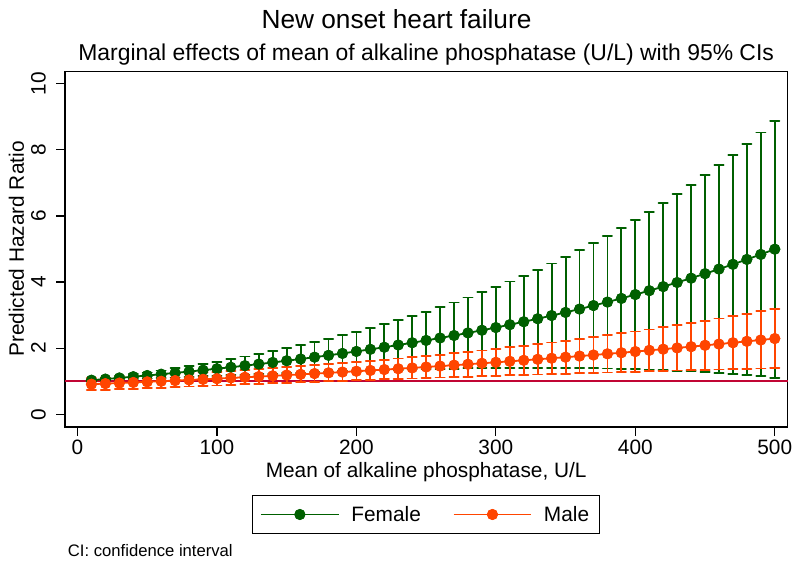

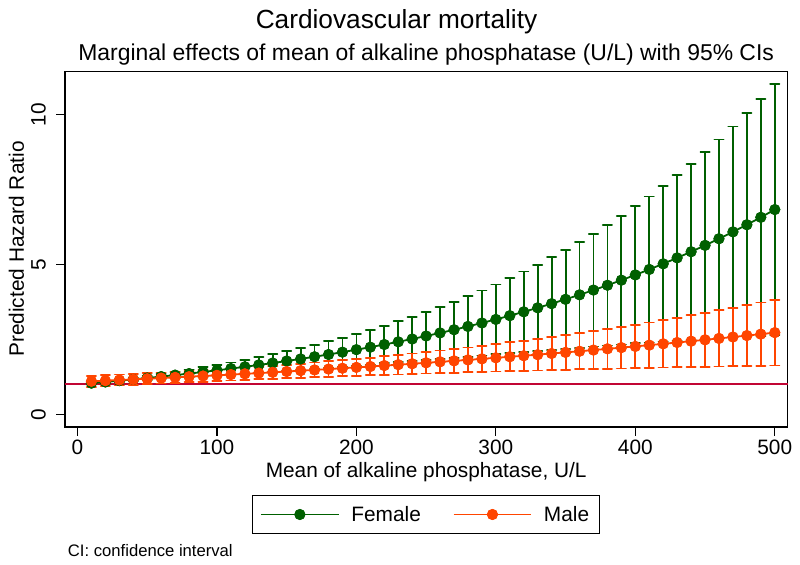

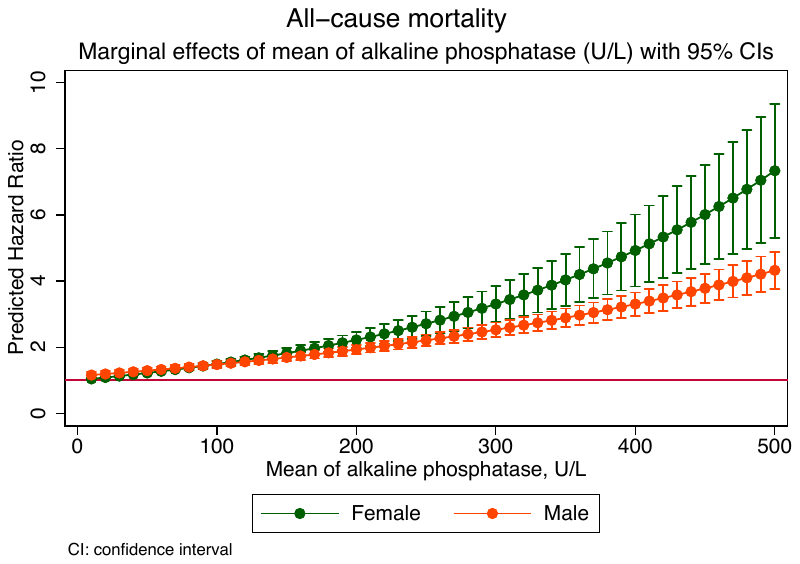

**Supplementary Figure 2B. Marginal effects of mean alkaline phosphatase stratified by gender to predict new onset heart failure, cardiovascular mortality, and all-cause mortality.**

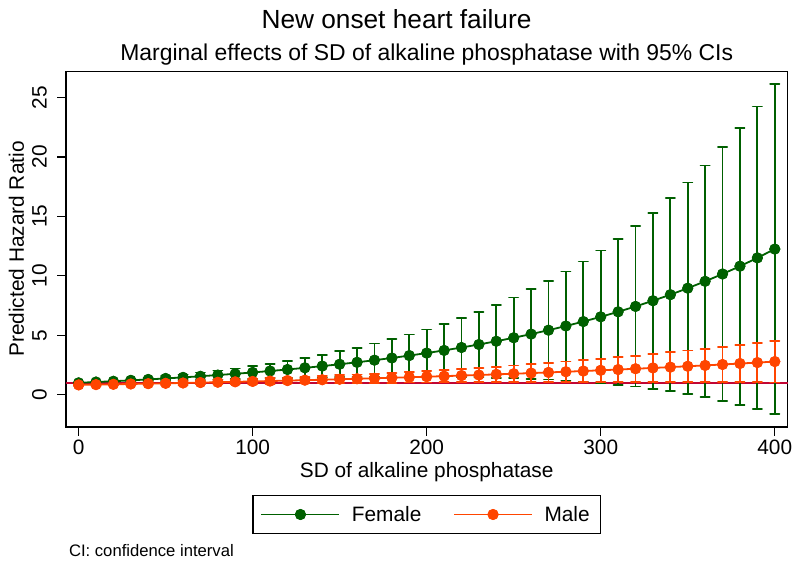

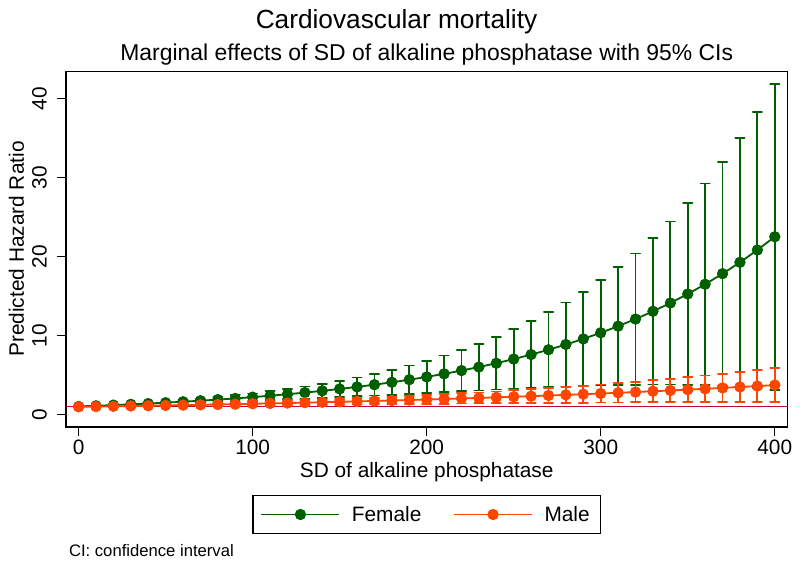

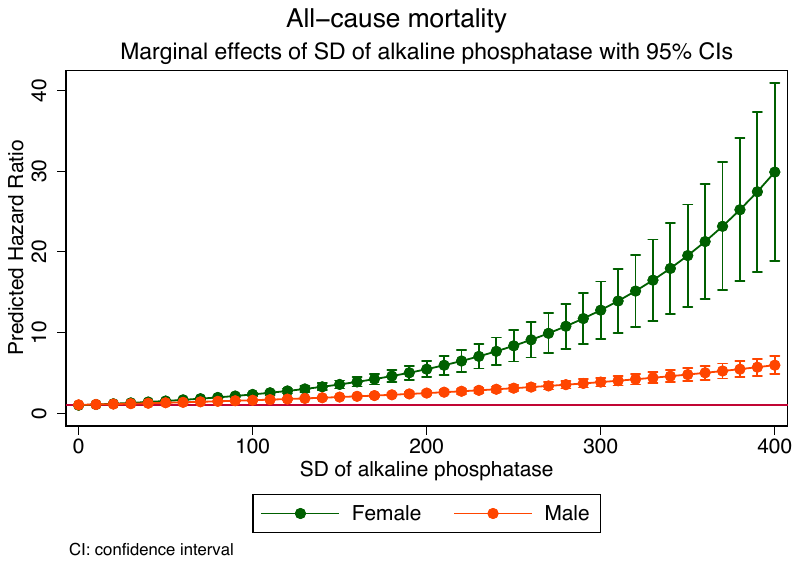

**Supplementary Figure 2C. Marginal effects of SD of baseline alkaline phosphatase stratified by gender to predict new onset heart failure, cardiovascular mortality, and all-cause mortality.**

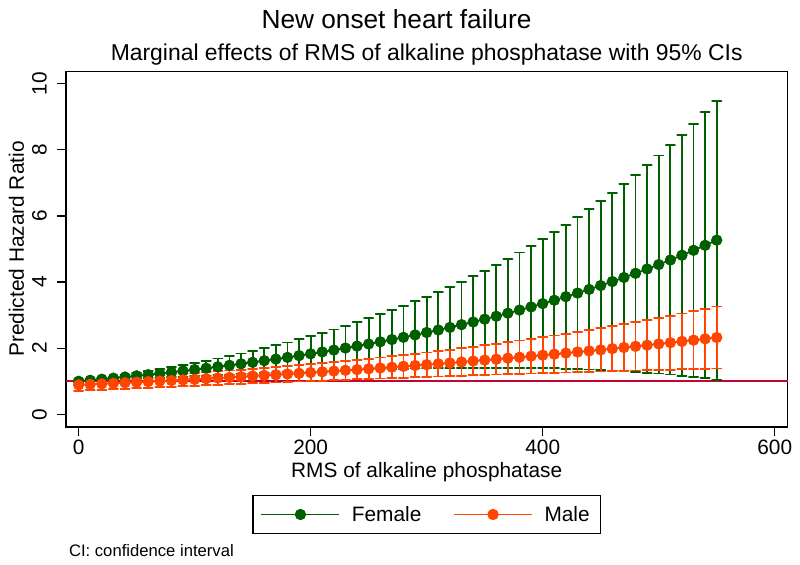

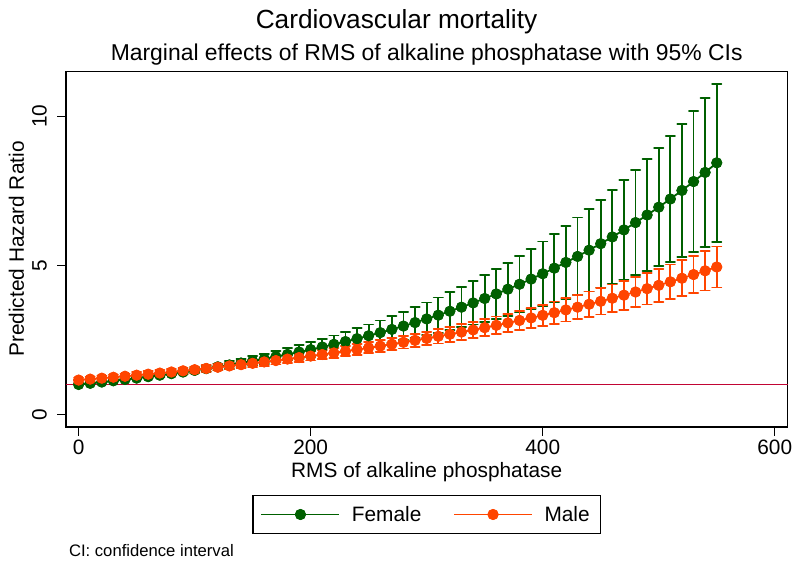

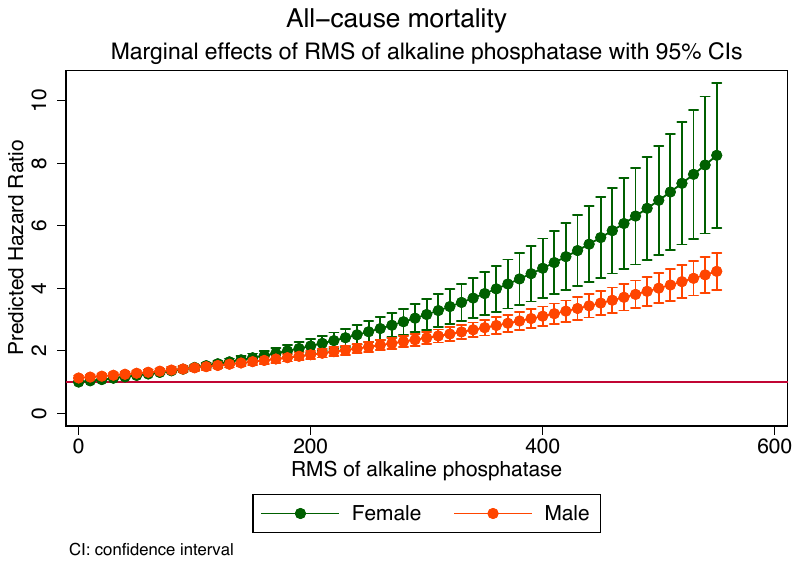

**Supplementary Figure 2D. Marginal effects of RMS of alkaline phosphatase stratified by gender to predict new onset heart failure, cardiovascular mortality, and all-cause mortality.**

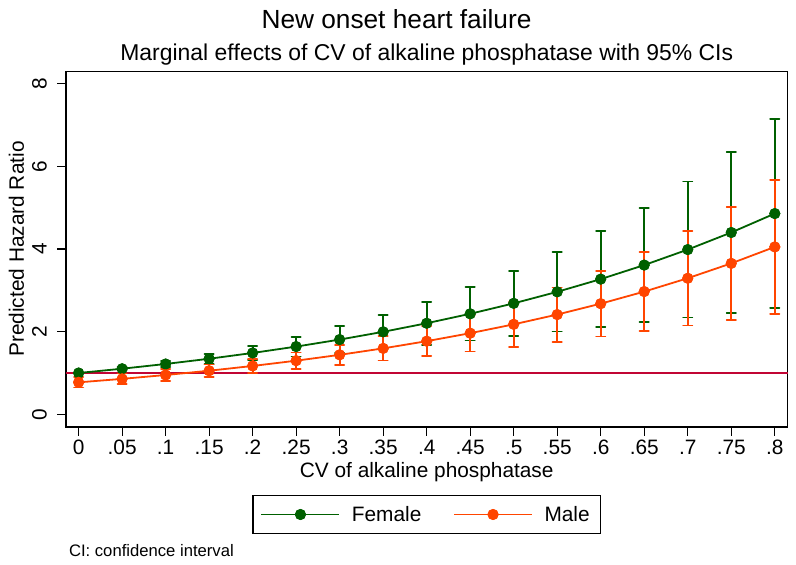

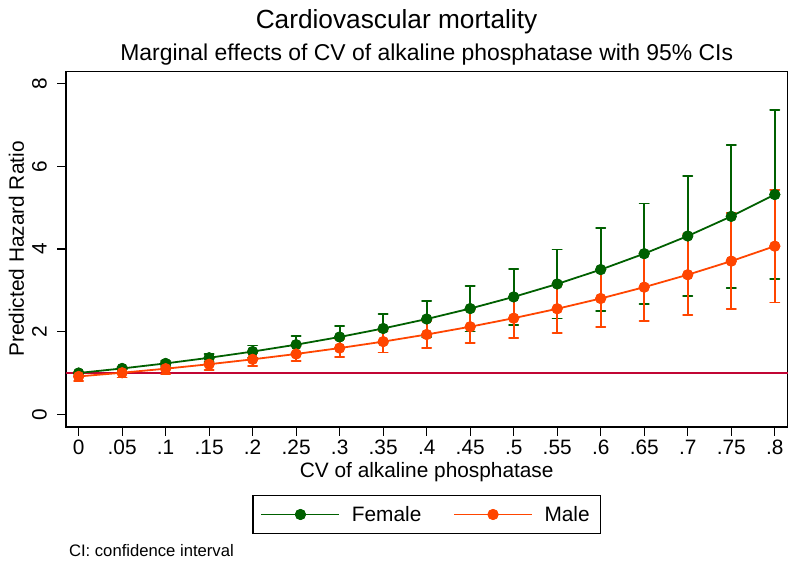

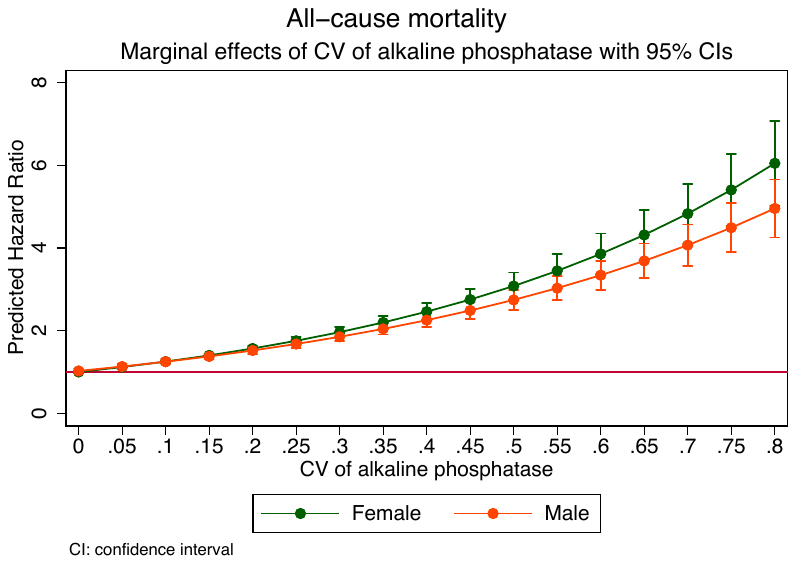

**Supplementary Figure 2E. Marginal effects of CV of alkaline phosphatase stratified by gender to predict new onset heart failure, cardiovascular mortality, and all-cause mortality.**

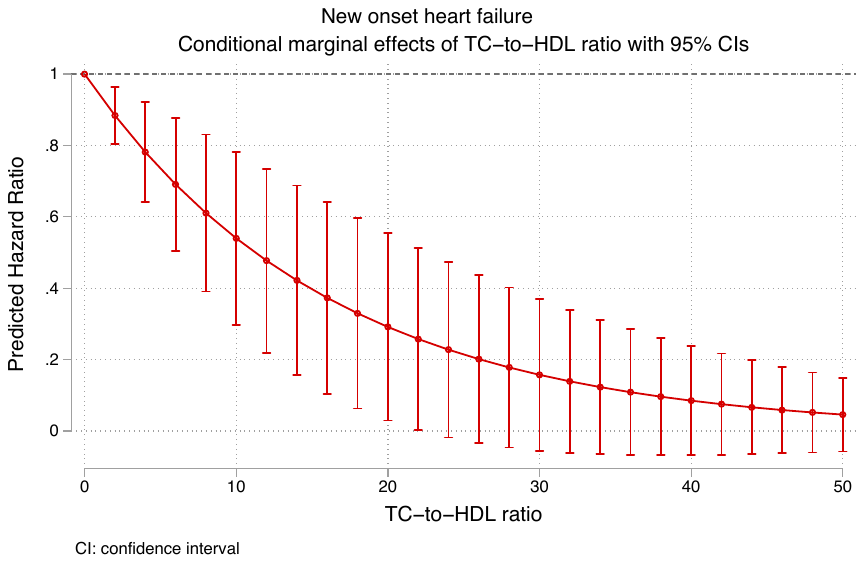

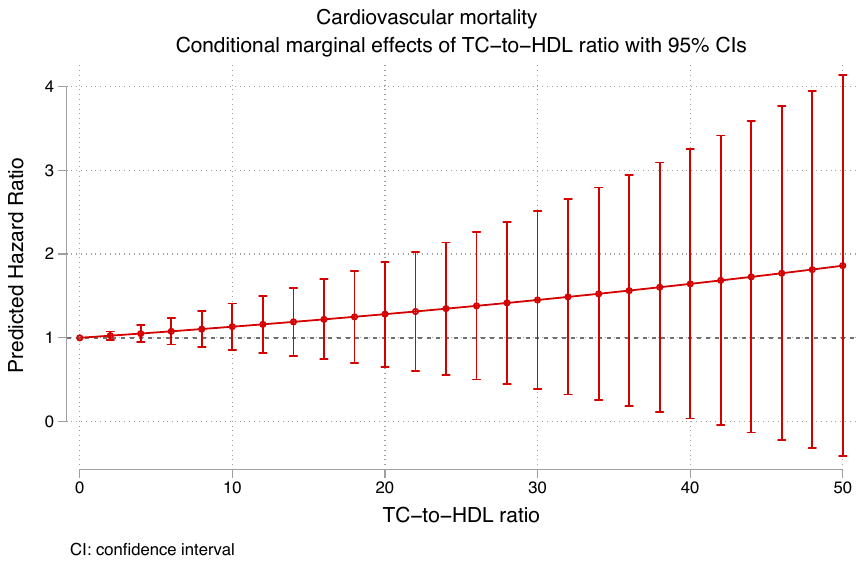

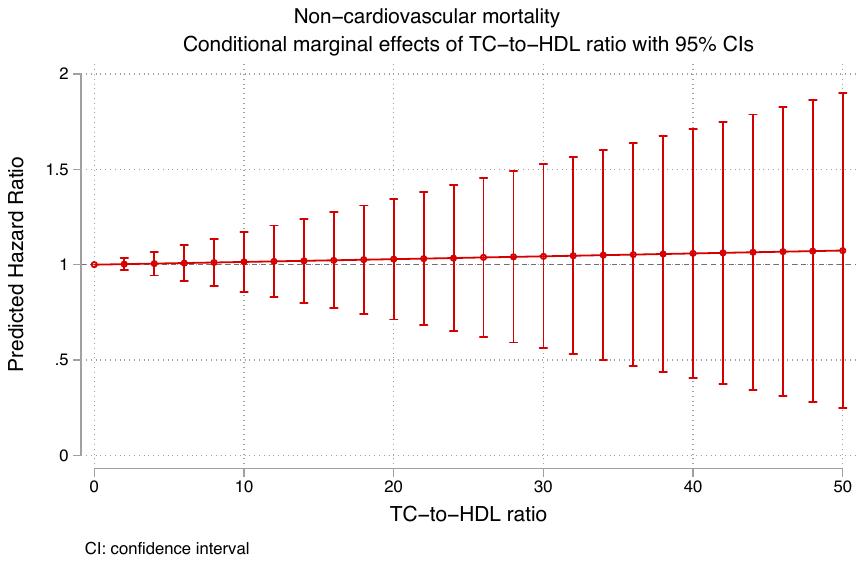

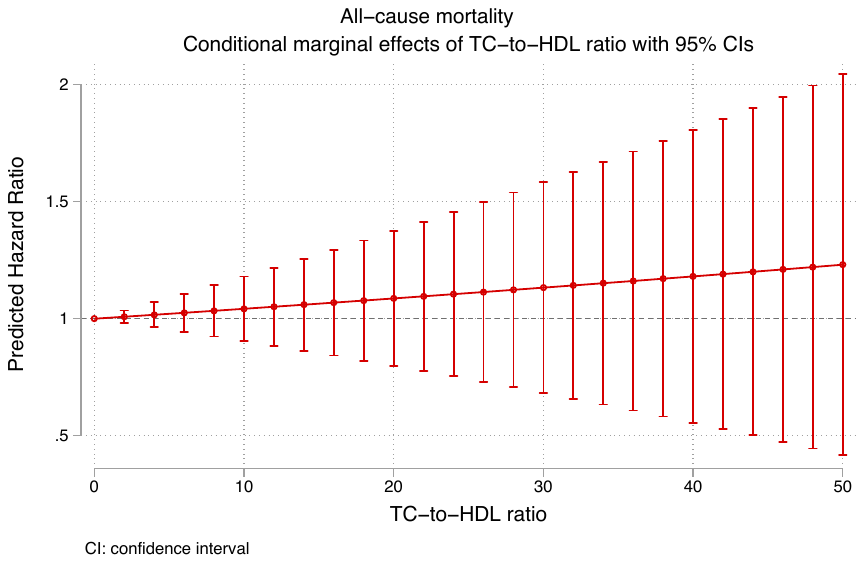

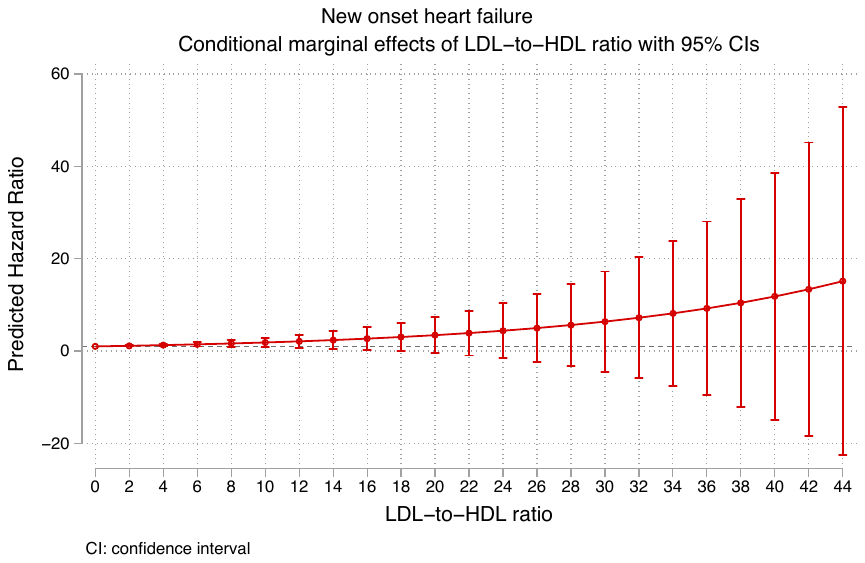

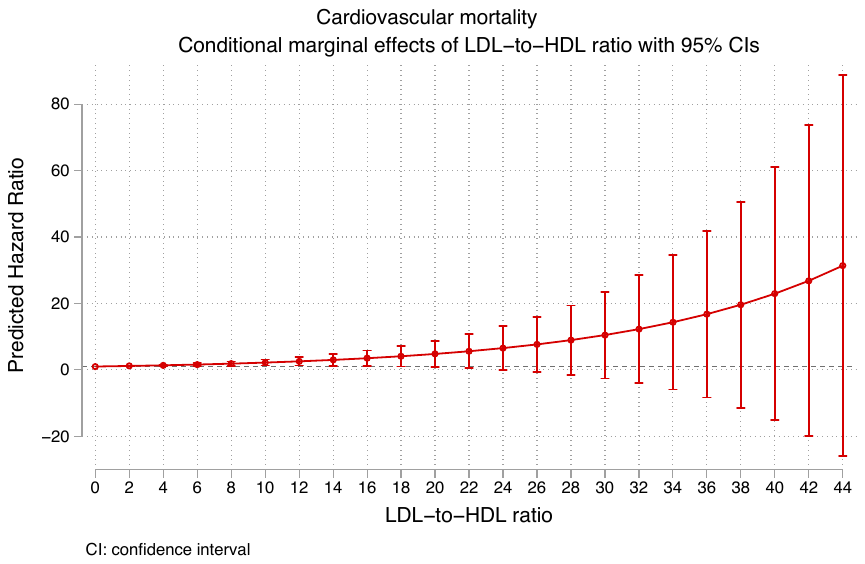

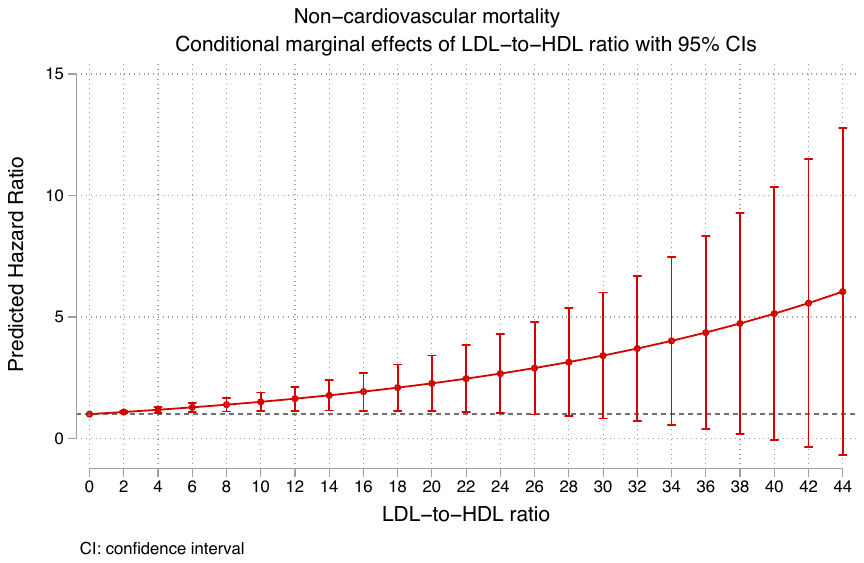

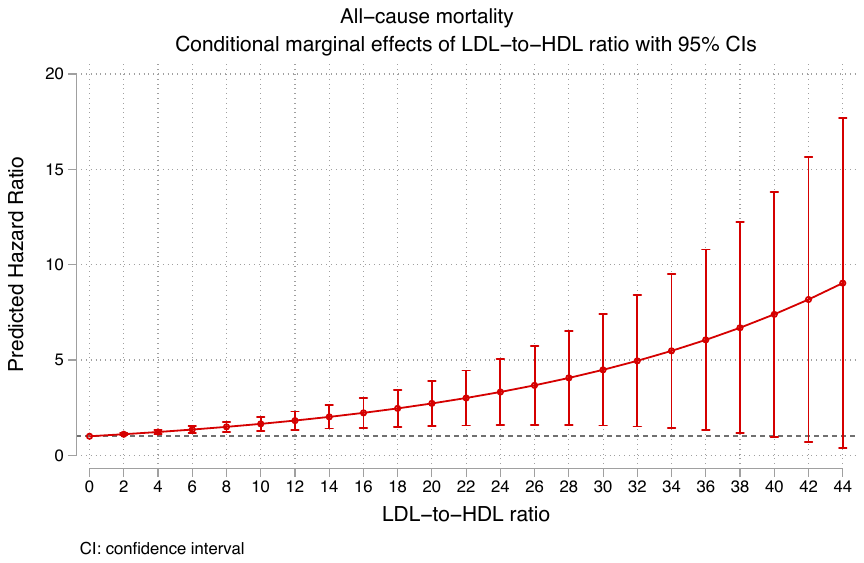

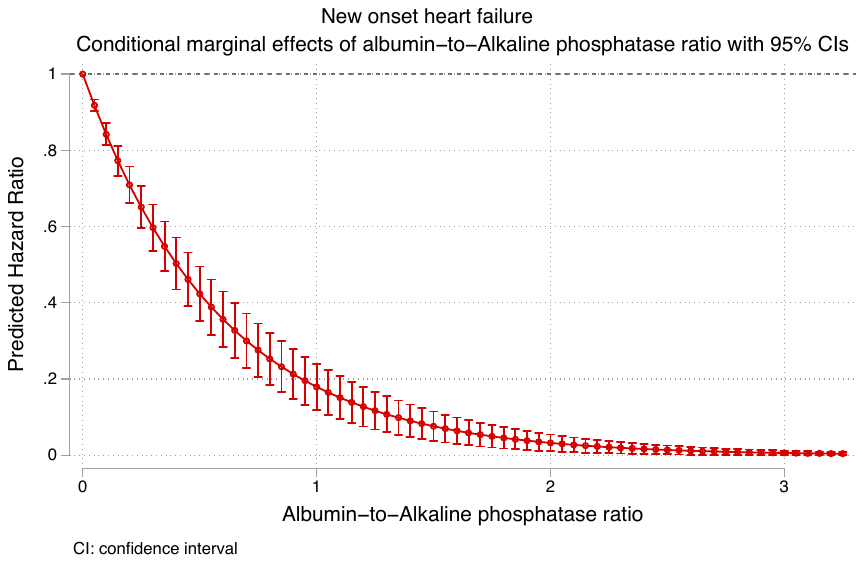

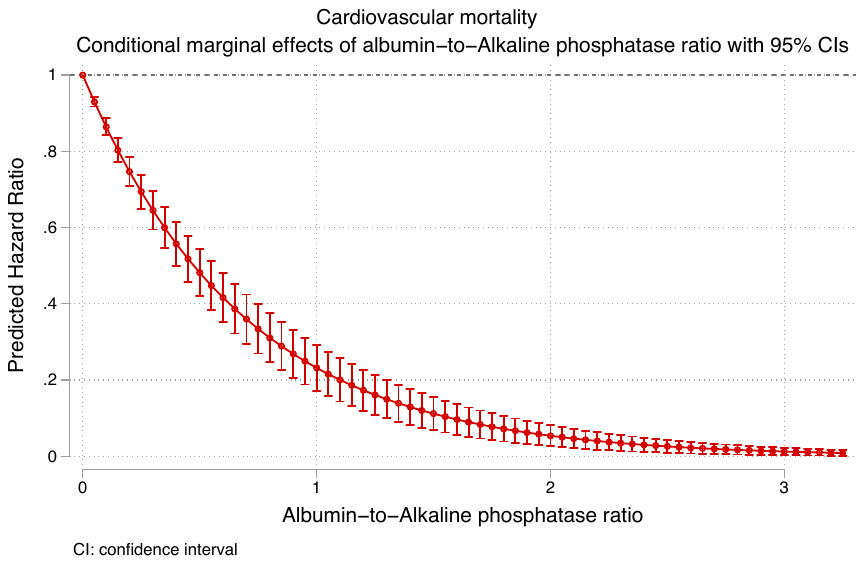

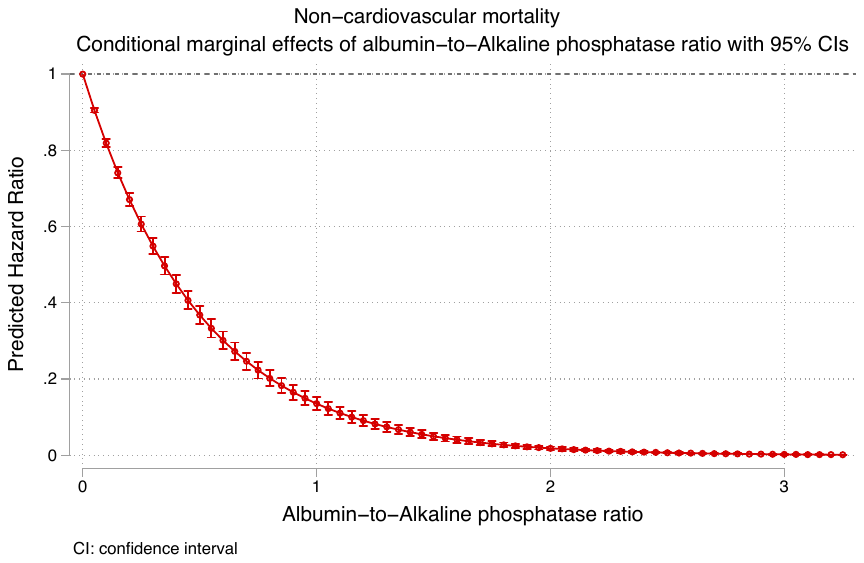

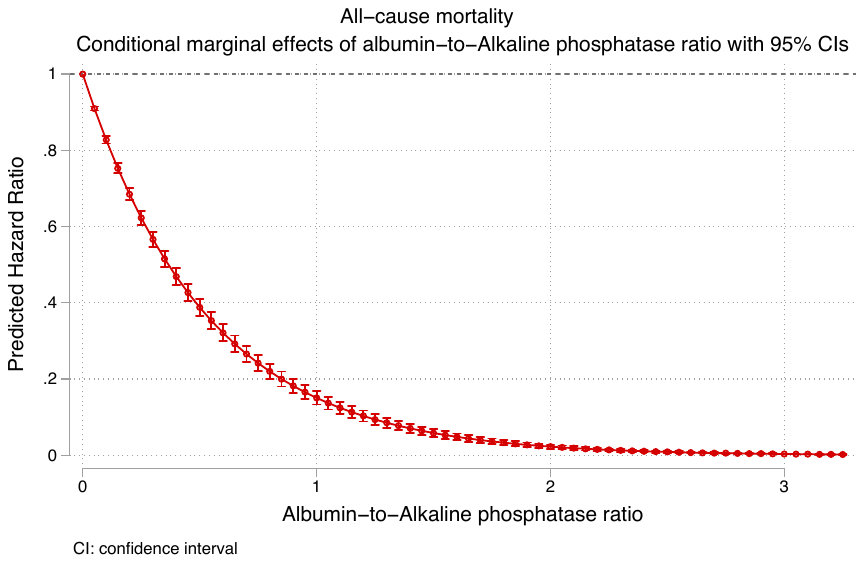

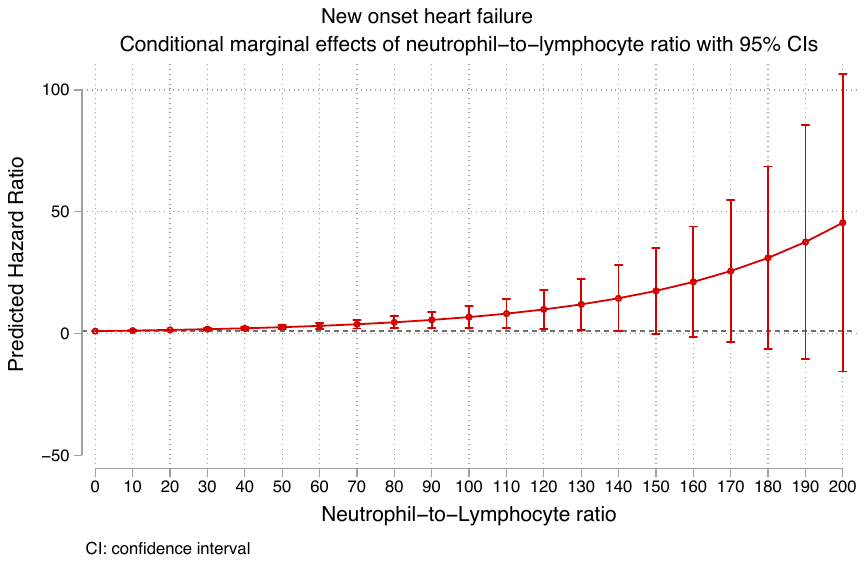

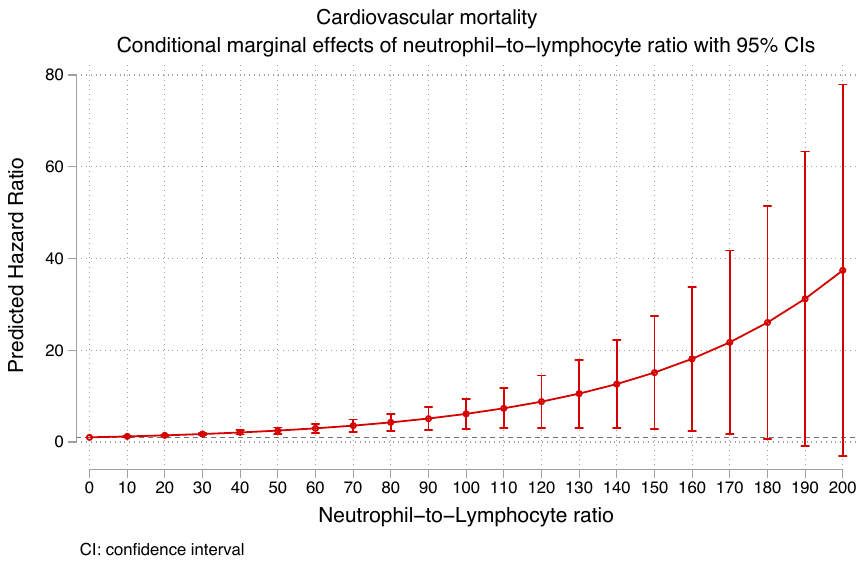

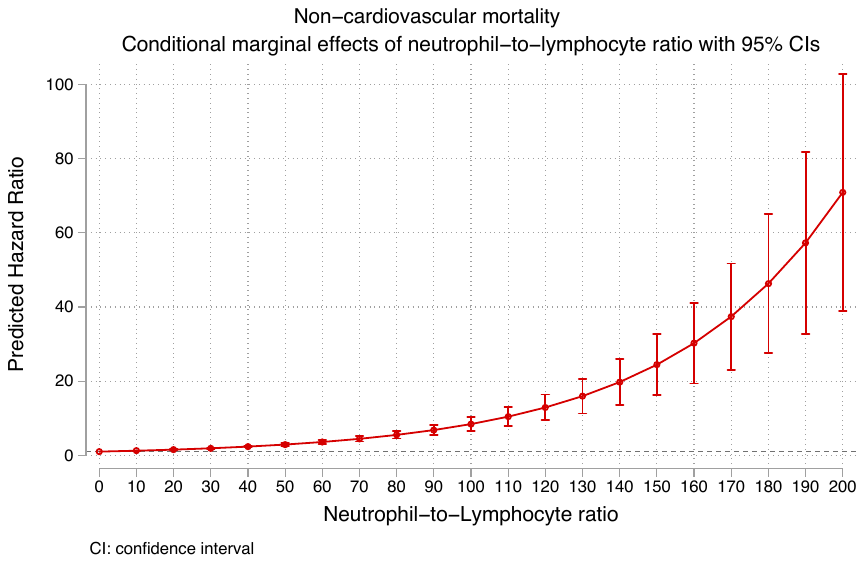

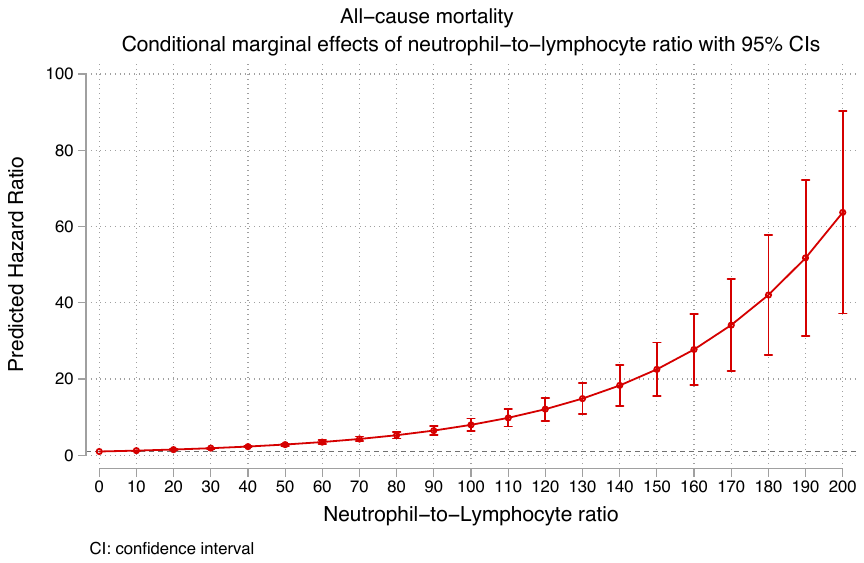

**Supplementary Figure 3. Marginal effects of subclinical inflammatory biomarkers to predict new onset heart failure, cardiovascular mortality, non-cardiovascular mortality, and all-cause mortality.**
